# Canine Rabies in N’Djaména: A Metapopulation SEIR Model Incorporating Vaccination and Inter-Patch Distances

**DOI:** 10.64898/2026.05.08.26352733

**Authors:** Hippolyte Djimramadji, Ousmane Koutou, Siguy Dawé

**Affiliations:** University of N’Djamena, Chad; CU-Kaya/Université Joseph KI-ZERBO, Burkina Faso

**Keywords:** Canine rabies, metapopulation model, SEIR model, vaccination, spatial heterogeneity, global stability

## Abstract

Canine rabies persists in N’Djaména (Chad) despite vaccination campaigns exceeding 70% coverage, suggesting a role for dog mobility and spatial heterogeneity. We propose a metapopulation SEIR model incorporating distance-modulated dog movements and an explicit vaccinated class. Analysis of the isolated patch establishes global stability of the disease-free equilibrium via a Lyapunov function. For the metapopulation, a composite Lyapunov function shows that elimination is governed by a reproduction number ℛ_*v*_. Calibrated with field data (2012–2022), simulations reveal that uniform vaccination of both patches reduces ℛ_*v*_ by 46% (from 2.84 to 1.52) but does not achieve elimination, while targeted strategies are less effective. These results demonstrate that exhaustive vaccination coverage across the entire urban network and increased vaccination intensity are necessary to eliminate canine rabies in N’Djaména. Our model provides a quantitative framework for planning effective control strategies.

## 1 Introduction

Canine rabies remains a major public health problem in sub-Saharan Africa, causing several thousand human deaths each year [9]. In N’Djaména, the capital of Chad, the canine population is estimated at around 23,600 dogs, most of which are owned but free-roaming, unvaccinated, and move freely throughout the city [10, 19]. Rabies transmission in dogs is the source of most human exposures [8].

To limit the spread of the disease, canine vaccination campaigns have been implemented, sometimes achieving over 70% vaccination coverage [12, 30]. These campaigns have temporarily interrupted rabies transmission, but sporadic resurgences have been observed, suggesting that the transmission dynamics are not fully understood [13].

Surveillance data provided by the *Institut de Recherche en Elèvage pour le Développement* ^1^ (IRED), Figures 2 to 4, allow us to trace the evolution of canine rabies in N’Djaména over the period 2012-2022. Figure 2 presents annual positive tests, revealing endemic persistence with high interannual variability. Monthly analysis (Figure 3) suggests a seasonal influence, with a concentration of cases in June-July. Finally, weekly data (Figure 4) show the fine dynamics of the infection, with successive epidemic outbreaks.

**Figure 1.**
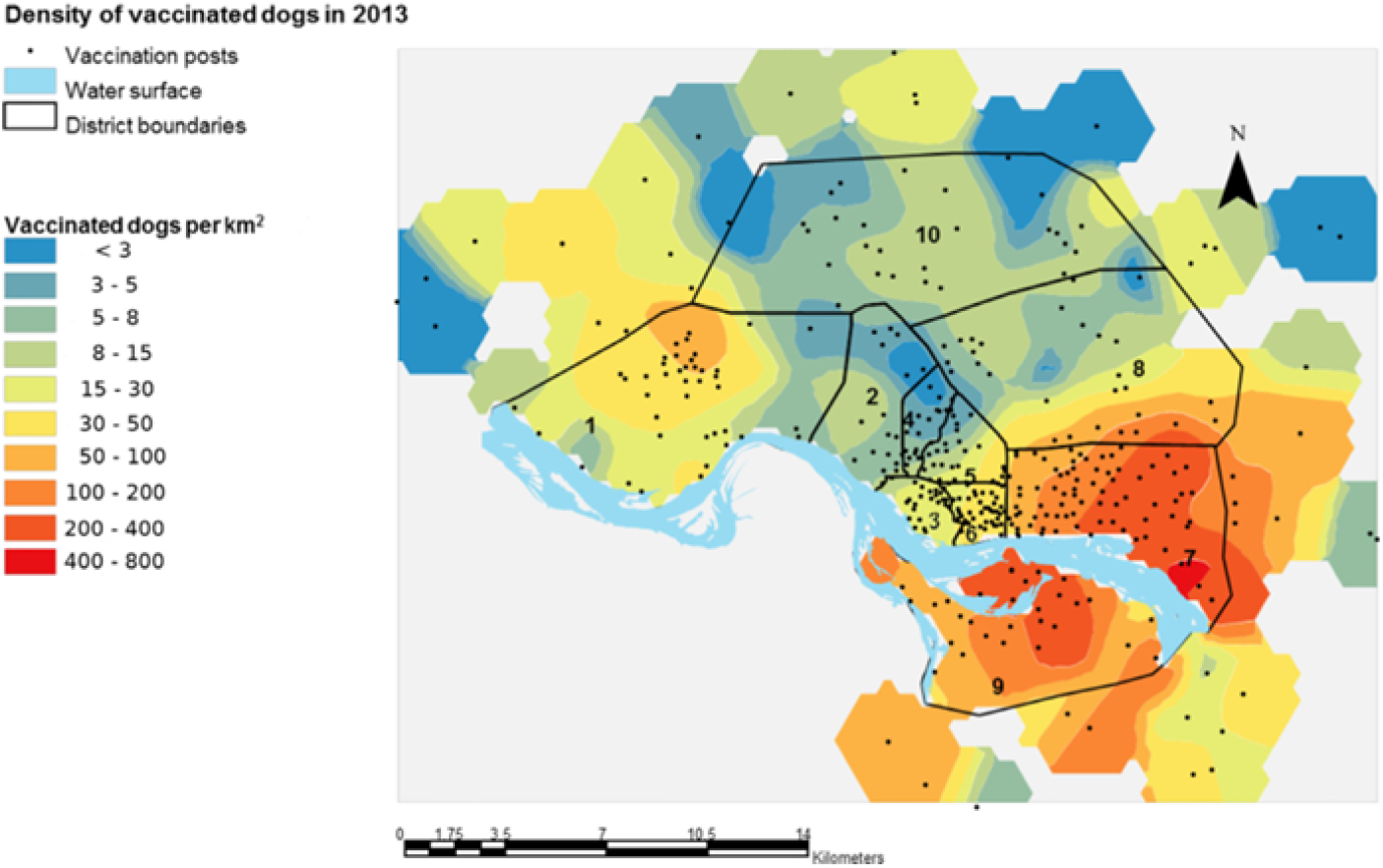
Districts of N’Djamena and density of vaccinated dogs in 2013, from [30]

**Figure 2.**
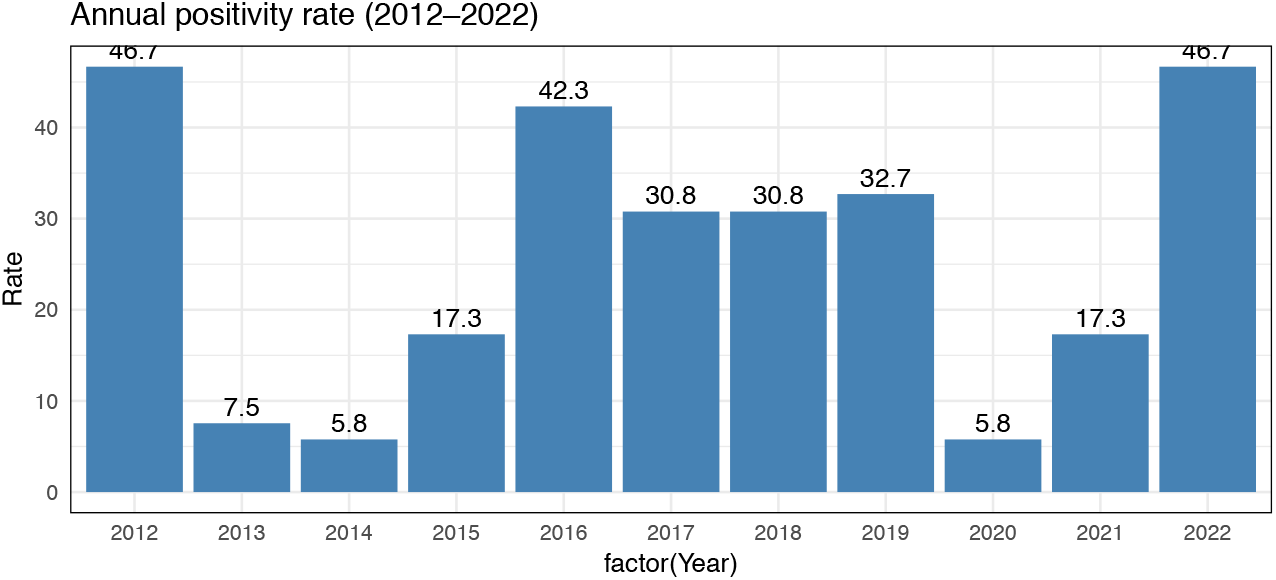
Annual tests from 2012-2022, source IRED.

**Figure 3.**
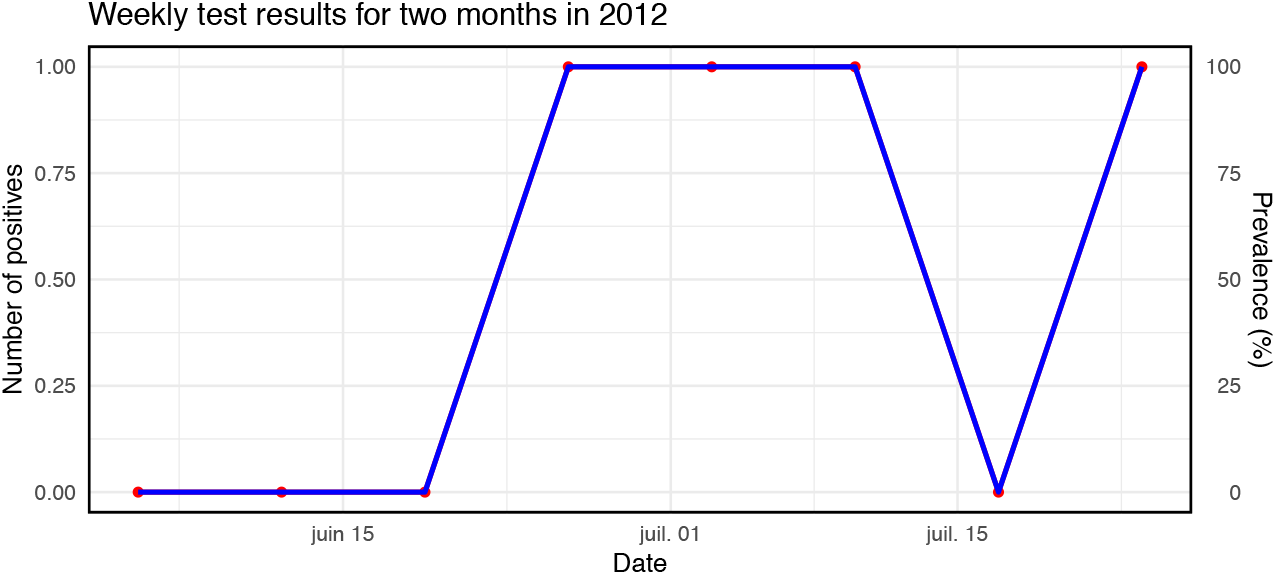
Positive tests from June-July 2012, source IRED.

**Figure 4.**
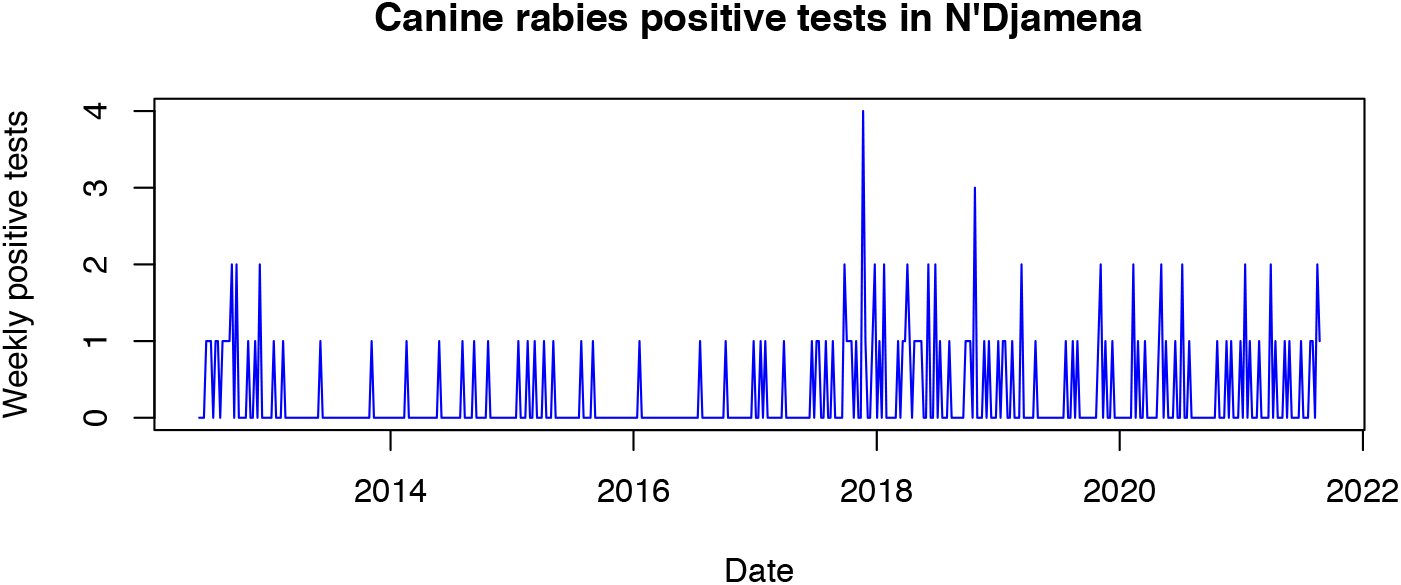
Weekly positive tests in N’Djamena from 2012-2022, source IRED.

Field studies conducted in N’Djamena between 2012 and 2018 showed that, despite campaigns achieving over 70% coverage, viral circulation persisted in certain peripheral districts [13, 16]. These observations suggest the existence of spatial heterogeneity in transmission, linked to dog mobility and disparities in vaccination coverage. Figure 1 illustrates the spatial heterogeneity of vaccination coverage across the districts of N’Djaména in 2013, with lower coverage in peripheral areas.

Complementary strategies such as canine population management (sterilization, vaccination, stray dog control) and optimization of vaccination center locations have been proposed to improve vaccination coverage and reduce transmission risk [4, 27]. Studying these dynamics is essential for designing effective and sustainable interventions against canine rabies and preventing associated human deaths.

Mathematical modeling has played a crucial role in understanding canine rabies dynamics and designing vaccination campaigns. Several approaches have been proposed, ranging from homogeneous compartmental models to individual-based and metapopulation models.

Several epidemiological models have been developed to study canine rabies: Laager et al. (2019) [13] proposed a metapopulation SEIR model for N’Djaména, incorporating dog movements between districts. *Limitation:* population flows do not explicitly account for distances between patches, and vaccination is not modeled. Colombi et al. (2020) [6] studied the spread of canine rabies at the national level in Africa, incorporating long-distance movements. *Limitation:* the model is calibrated at the national rather than urban scale, limiting its applicability to the specific dynamics of N’Djaména. Sararat et al. (2022) [23] developed a stochastic individual-based model, taking into account the spatial structure of buildings and roads. *Limitation:* the model is applied to Asian cities and does not distinguish vaccinated dogs. Thus, although several studies have contributed to understanding transmission mechanisms, the joint consideration of spatial distance between patches and targeted vaccination remains underexplored, particularly in the context of N’Djaména. More recent research has focused on the social and institutional determinants of rabies control in N’Djamena. A qualitative study conducted by [15] highlighted the barriers to canine vaccination at the community and institutional levels, including lack of awareness, poor accessibility of vaccination points, and persistent mistrust among some dog owners. These results underscore the importance of integrating socio-behavioral dimensions into rabies control strategies, in addition to purely epidemiological approaches.

Furthermore, an ongoing research project (2021–2024) entitled *Multiscale Transmission Dynamics of Rabies in Africa: The Urban-Rural Interface* [25] focuses on the multiscale dynamics of rabies in Africa, with a particular emphasis on the urban-rural interface in N’Djamena. This project aims to understand how dog movements between urban and rural areas influence disease persistence, and to develop models integrating both spatial dimensions and socio-economic interactions. These recent works enrich the understanding of canine rabies in N’Djamena and confirm the need for an integrated approach combining mathematical modeling, social sciences, and participatory strategies. Our model fits within this perspective by seeking to quantify the impact of spatial distance and targeted vaccination coverage on rabies dynamics in the Chadian capital. [30] showed that mass dog vaccination in N’Djaména (71% coverage) temporarily interrupts rabies transmission, but genetic analysis revealed reintroductions from outside the city after nine months. This finding suggests that spatial heterogeneity and dog mobility drive disease persistence, a hypothesis we test quantitatively. While [30] documented the *existence* of reintroductions, our model determines the *threshold conditions* under which uniform versus targeted vaccination can achieve elimination.

In this context, recent field observations and socio-behavioral analyses confirm that rabies dynamics in N’Djaména are influenced not only by epidemiological interactions between dogs but also by urban mobility and disparities in vaccination coverage. To better understand and quantify these effects, we propose an SEIR model, inspired by the work of [13] and [30], in a metapopulation explicitly incorporating inter-patch flows modulated by distance and a distinct class of vaccinated dogs. Our objective is to evaluate how distance between patches and targeted vaccination coverage influence the persistence or disappearance of rabies, and then to explore the most effective control strategies in the specific context of N’Djaména.

## 2 Model description and comparison with previous work

We consider N’Djaména as a set of interconnected *patches* (districts or peripheral villages). The canine population in each patch is divided into five classes: susceptible (*S*_*i*_), exposed (*E*_*i*_), infectious (*I*_*i*_), removed (*R*_*i*_), and vaccinated (*V*_*i*_).

Unlike previous models [6, 13, 23]:

1. We introduce an explicit class of vaccinated dogs (*V*_*i*_), allowing us to track immunized dogs and evaluate campaign effectiveness.
2. Inter-patch flows are modulated by the distance between patches (*f* (*d*_*ij*_)), reflecting the actual probability of dog movement.
3. Vaccination is spatially targeted based on distance to the campaign center (*g*(*d*_*iC*_)) (Figure 1), which was not considered in previous work.

The flow *γ*_*i*_*I*_*i*_ to *R*_*i*_ represents the removal of infectious dogs, including capture, isolation, or euthanasia. This compartment *does not mean biological recovery*, as rabies is almost always fatal once symptoms appear. For exposed dogs (*E*_*i*_), early treatment (post-exposure vaccination or immunoglobulins) is considered via a possible flow to *R*_*i*_ or *V*_*i*_, reducing the probability of progression to *I*_*i*_ [9, 13]. Dogs can move between patches, modulated by the distance *d*_*ij*_ via the function 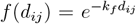. Vaccination is spatially targeted based on distance to the campaign center *d*_*iC*_, represented by 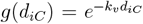. These exponential functions reflect the idea that dog movements and campaign effectiveness decrease rapidly with distance.

## 3 Mathematical formulation

Inter-patch flows and vaccination are modulated by distance to reflect the reality of dog movements and access to vaccination centers. More specifically, the dog movement flow between patches *i* and *j* is weighted by the decreasing function 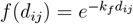, which is consistent with metapopulation approaches used in canine rabies modeling [9, 13]. Similarly, the probability that a dog in patch *i* is vaccinated decreases with distance to the campaign center *C*, represented by 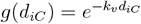, following principles similar to targeted vaccination in urban contexts [26, 29].

- Λ_*i*_: dog entry rate into patch *i* (birth or immigration) [dog/day]
- *β*_*i*_: rabies transmission rate in patch *i* [1/(dog·day)]
- *µ*_*i*_: natural mortality rate of dogs in patch *i* [1/day]
- *α*_*i*_: rabies-induced mortality rate in patch *i* [1/day]
- *σ*_*i*_: rate at which exposed dogs become infectious in patch *i* [1/day]
- *γ*_*i*_: removal rate of infectious dogs (capture, isolation, or euthanasia) or recovery rate in patch *i* [1/day]
- *v*_0_: maximum vaccination rate at the campaign center [1/day]
- *d*_*iC*_: distance between patch *i* and the vaccination center *C* [km]. This distance influences the probability of vaccination for dogs in patch *i*.
- 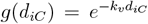: spatial decay function for vaccination. It represents the probability that a dog in patch *i* is vaccinated based on its distance from the center.
- *k*_*v*_: spatial decay coefficient for vaccination [1/km], controlling how quickly campaign effectiveness decreases with distance *d*_*iC*_.
- 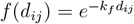: decay function for dog movement flow between patches *i* and *j* based on distance *d*_*ij*_ [km]. It represents the probability that a dog moves from patch *i* to patch *j*.
- *k*_*f*_ : spatial decay coefficient for flows [1/km]. The larger *k*_*f*_, the more movements are limited to short distances.
- *d*_*ij*_: distance between patch *i* and patch *j* [km].
- *m*_0_: baseline dog movement rate between patches [1/day]
- *N*_*i*_ = *S*_*i*_ + *E*_*i*_ + *I*_*i*_ + *R*_*i*_ + *V*_*i*_: total dog population in patch *i* [dog]

For each patch *i* = 1, 2, …, *n*, the differential equations governing canine population dynamics from figure 5:

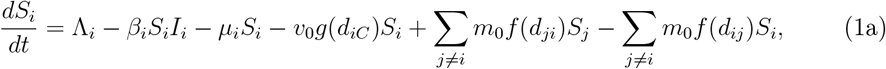

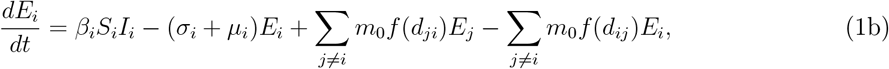

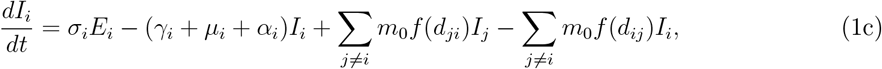

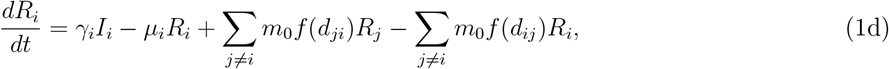

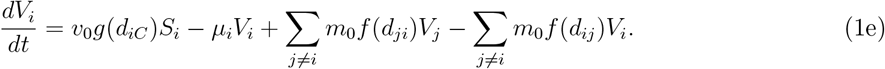

**Figure 5.**
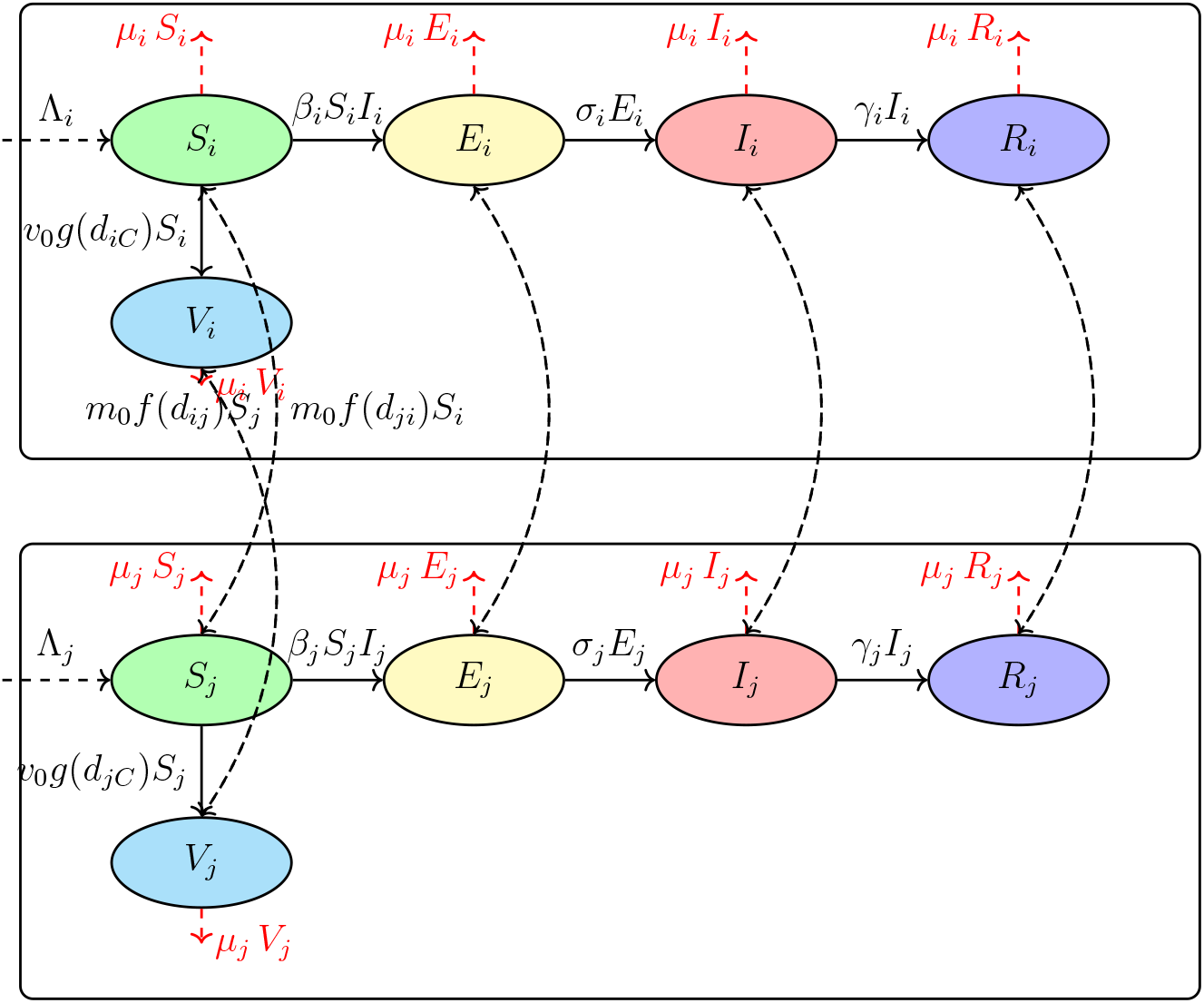
Flow diagram of the canine rabies model between two patches *i* and *j*.

The total population of patch *i* is given by:

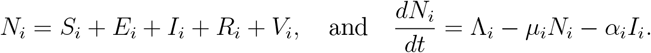

## 4 The isolated model in one (patch)

Consider the isolated model representing the dynamics of rabies in a closed canine population, denoted *i*. Let *S*_*i*_(*t*), *E*_*i*_(*t*), *I*_*i*_(*t*) and *R*_*i*_(*t*) be respectively the populations of *susceptible, exposed, infectious* and *removed* (immune or dead from the disease) dogs at time *t*. The model is written as follows:

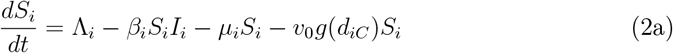

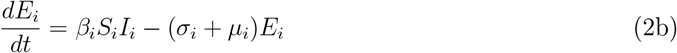

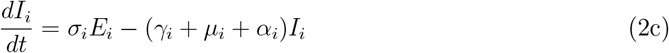

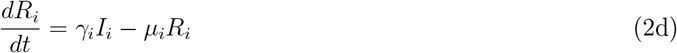

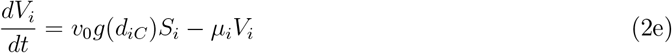

Let *X*_*i*_ = (*S*_*i*_, *E*_*i*_, *I*_*i*_, *R*_*i*_, *V*_*i*_)^*T*^.

### 4.1 Analysis of the isolated model

#### Lemma 1.

*The set*

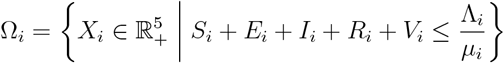

*is positively invariant under the flow of* (2), *meaning that any trajectory starting from* Ω_*i*_ *remains in* Ω_*i*_ *for all t* > 0.

*Proof*. The system (2) is of class 𝒞 ^1^ on 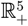, thus locally Lipschitz. We rewrite the equations of (2) in the form

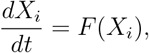

with *F* (*X*_*i*_) locally Lipschitz on 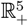, and for each component, outgoing flows are proportional to the corresponding variable. According to the *Cauchy–Lipschitz (or Cauchy–Picard) theorem* ([21, Theorem 1.1]), there exists a unique solution *X*_*i*_(*t*) to system (2). Thus, if 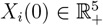, then 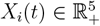, for all *t* > 0, meaning that the variables remain positive.

Moreover, let

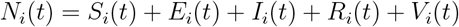

be the total population of patch *i*.

By adding the equations of (2), we obtain:

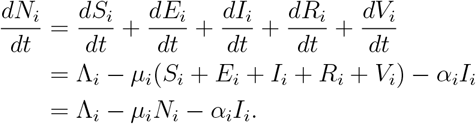

Since *α*_*i*_*I*_*i*_ ≥ 0, then

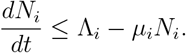

The solution is:

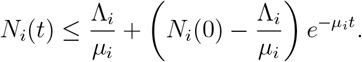

Thus,

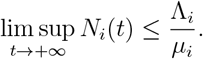

Furthermore, if 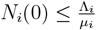, we have 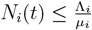 for all *t* > 0.

#### 4.1.1 Analysis of the isolated model without vaccination

In the absence of vaccination, i.e., when *V*_*i*_ = 0, the system reduces to:

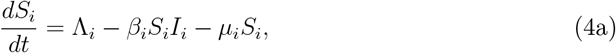

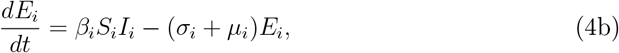

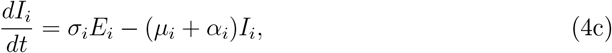

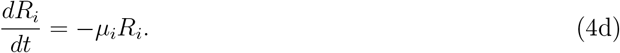

Disease-free equilibrium (DFE): by setting the derivatives to zero in (4) and assuming *E*_*i*_ = *I*_*i*_ = *R*_*i*_ = 0, we obtain:

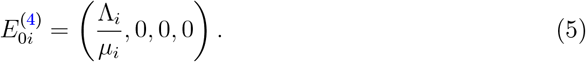

Basic reproduction number: applying the *next generation matrix* method from [28], we obtain:

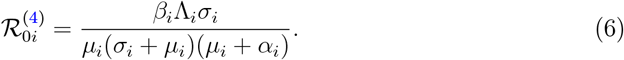

The endemic equilibrium of system (4) is written as:

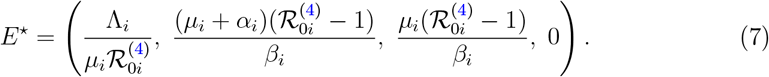

The disease-free equilibrium 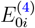 is locally asymptotically stable if 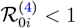, and unstable if 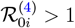.

Thus, the condition 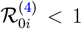 guarantees that any introduction of infectious individuals into the population does not lead to an epidemic in the system.

#### 4.1.2 Analysis of the isolated model with vaccination

The system admits a disease-free equilibrium (DFE) obtained by setting *E*_*i*_ = *I*_*i*_ = 0:

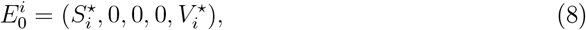

where

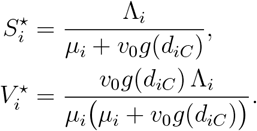

The basic reproduction number associated with site *i* is obtained from the new infection matrix *F* and the transition matrix *V*.

Taking (*E*_*i*_, *I*_*i*_)^⊤^, the functions associated with new infections ℱ (*E*_*i*_, *I*_*i*_) and transitions 𝒱 (*E*_*i*_, *I*_*i*_) are

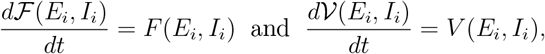

where, at the disease-free equilibrium 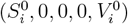:

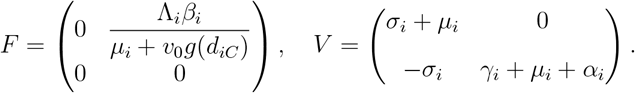

The inverse of matrix *V* is given by:

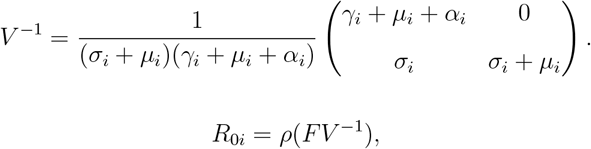

where *ρ*(·) denotes the spectral radius. For (2), we have:

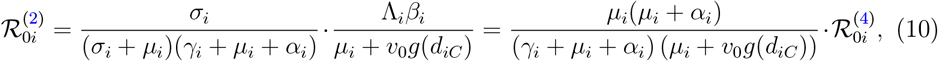

with 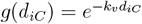 the spatial decay function for vaccination.

When 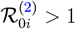, system (2) admits an endemic equilibrium 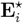 such that 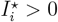 given by

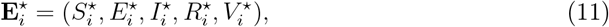

where

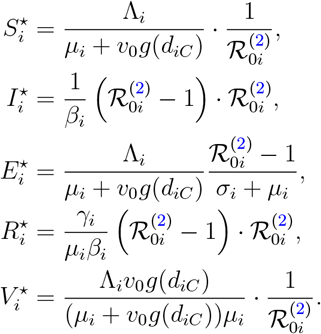

### 4.2 Local and global stability analysis of the DFE

#### Theorem 2.

*The DFE* 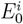 *of system* (2) *is locally asymptotically stable if* 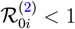 *and unstable if* 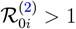.

*Proof*. To study the local stability of the DFE 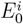, we linearize system (2) around 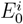. Since the infectious variables are (*E*_*i*_, *I*_*i*_), the linearized dynamics of the infectious subsystem are:

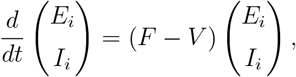

where *F* and *V* are the new infection and transition matrices given earlier:

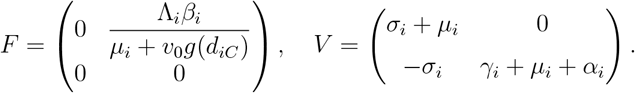

The basic reproduction number is

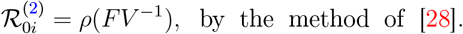

The eigenvalues of the linearized system are the roots of the characteristic polynomial of *V* − *F*, given by:

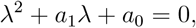

with

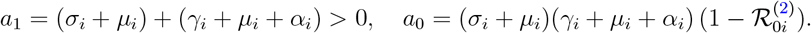

By the Routh–Hurwitz criterion [22], both roots of this polynomial have negative real parts if and only if *a*_1_ > 0 and *a*_0_ > 0, i.e., when 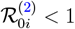.

Thus, the DFE 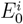 is locally asymptotically stable if 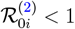, and unstable otherwise.

#### Theorem 3.

*The DFE* 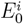 *of system* (2) *is globally asymptotically stable in* Ω_*i*_ *if* 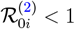.

*Proof*. We rewrite system (2) in the form:

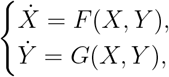

where *X* = (*S*_*i*_, *R*_*i*_, *V*_*i*_) corresponds to the non-infectious classes and *Y* = (*E*_*i*_, *I*_*i*_) to the infectious classes.

When *Y* = 0, the subsystem 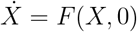 reduces to:

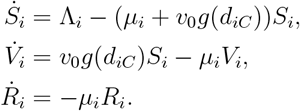

This subsystem is linear and admits a globally asymptotically stable equilibrium

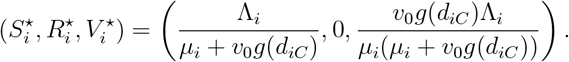

The infectious part can be written as:

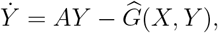

where

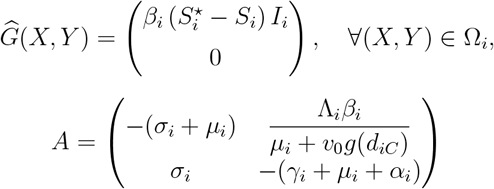

is a Metzler matrix (off-diagonal entries non-negative) and 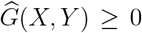 for all (*X, Y*) ∈ Ω_*i*_.

Thus, conditions (C1) and (C2) of [2] are satisfied. Therefore, according to the theorem of Castillo-Chavez and Song [3], if 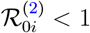, the DFE

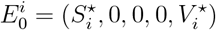

is globally asymptotically stable in Ω_*i*_.

We propose an alternative proof of the global stability of the disease-free equilibrium (DFE) for the isolated model (2), by constructing an appropriate Lyapunov function. This approach, inspired by the work of [24, 17], allows us to establish global stability without relying on the Castillo-Chavez and Song theorem.

#### Theorem 4.

*If* 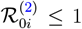 *then the disease-free equilibrium* 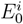 *of system* (2) *is globally asymptotically stable in the positively invariant set* Ω_*i*_ *defined by:*

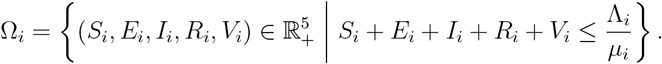

*Proof*. Consider the following Lyapunov function, defined on Ω_*i*_:

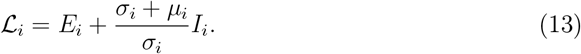

This function is positive definite with respect to the equilibrium 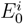: it vanishes when *E*_*i*_ = *I*_*i*_ = 0 and is strictly positive elsewhere.

Let us compute the derivative of ℒ_*i*_ along the trajectories of system (2). Using equations (2c) and (2b), we obtain:

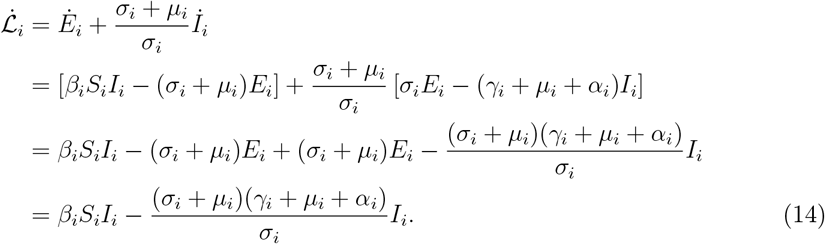

Thus:

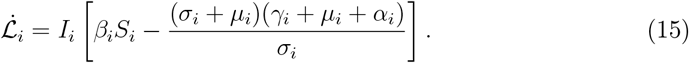

In the invariant set Ω_*i*_, we have 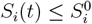 for all *t* ≥ 0. Indeed, according to Lemma 1, the total population *N*_*i*_(*t*) is bounded by Λ_*i*_*/µ*_*i*_, and equation (4a) shows that *S*_*i*_ is bounded by its equilibrium value 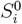 when *I*_*i*_ = 0; a more refined analysis using differential comparison establishes that 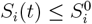 for any initial condition in Ω_*i*_. Consequently:

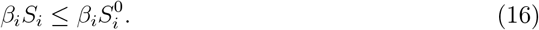

Substituting (16) into (15), we obtain:

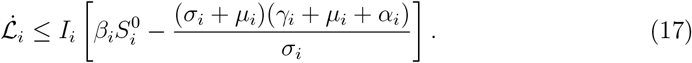

Recalling the expression for the basic reproduction number of the isolated model with vaccination (10), we deduce:

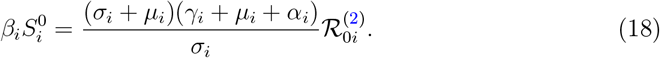

Substituting (18) into inequality (17), we obtain:

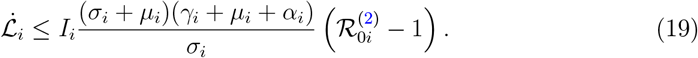

Case 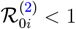: then 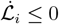 for all (*S*_*i*_, *E*_*i*_, *I*_*i*_, *R*_*i*_, *V*_*i*_) ∈ Ω_*i*_, with 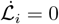 if and only if *I*_*i*_ = 0. The largest invariant set contained in 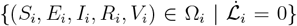 is the set where *I*_*i*_ = 0. Setting *I*_*i*_ = 0 in system (2), we obtain *E*_*i*_ → 0 (since 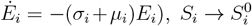 and 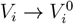 according to equations (4a) and (2e). Thus, the only invariant point in this set is the DFE 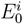. By LaSalle’s invariance principle [14], every trajectory starting from Ω_*i*_ converges to 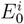. Hence, the equilibrium 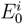 is globally asymptotically stable.

Case 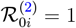: then 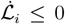 with equality when *I*_*i*_ = 0. The same LaSalle argument shows convergence to 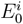.

Case 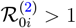: in a neighborhood of the DFE, we have 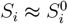 and 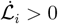 for sufficiently small *I*_*i*_ > 0. Thus, the DFE is unstable.

Therefore, we have established that the DFE 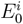 is globally asymptotically stable in Ω_*i*_ when 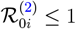, and unstable when 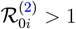.

#### Remark 5.

*The Lyapunov function* (13) *is a linear combination of the infected classes E*_*i*_ *and I*_*i*_, *with coefficients chosen so that the interaction terms between E*_*i*_ *and I*_*i*_ *cancel out. This construction is standard for SEIR-type models [24]. The extension to the metapopulation model* (20) *can be achieved by considering a composite Lyapunov function:*

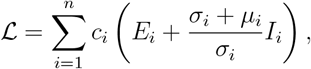

*where the coefficients c*_*i*_ *are the components of the left eigenvector associated with the transition matrix V of the metapopulation system, according to the method developed in [24] for coupled systems*.

### 4.3 Sensitivity and elasticity analysis of parameters

A local sensitivity analysis of 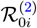 with respect to the model parameters helps identify those that most influence infection transmission within site *i*. Elasticity indices are defined by:

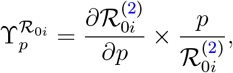

where *p* denotes a model parameter.

For the isolated model (2), the elasticity indices of 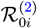 with respect to each parameter are:

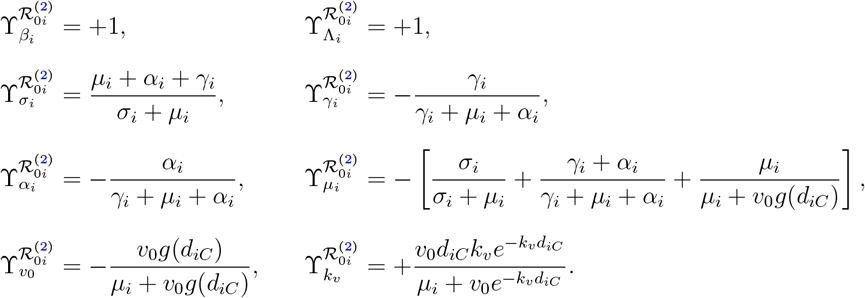

Thus

- parameters *β*_*i*_, Λ_*i*_ and *σ*_*i*_ increase 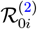 when they increase;
- while *µ*_*i*_, *γ*_*i*_, *α*_*i*_ and *v*_0_ reduce it.

This analysis thus helps guide the most effective control and vaccination strategies for patch *i*.

## 5 Analysis of the movement model (metapopulation)

We now consider the complete system where the *n* patches (districts of N’Djaména) are interconnected by dog movements.

### 5.1 Vector formulation of the metapopulation model

Let the state vectors of dimension *n* be:

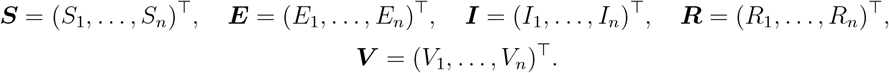

We also define the diagonal matrices of parameters:

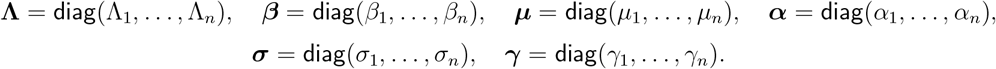

The spatial modulation function for vaccination is encapsulated in the diagonal matrix

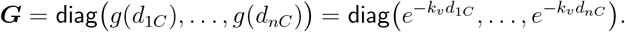

Finally, the movement matrices ℳ_*S*_, ℳ_*E*_, ℳ_*I*_, ℳ_*R*_, ℳ_*V*_ are constructed from the movement rates 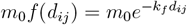. For *X* ∈ *{S, E, I, R, V*}, we have:

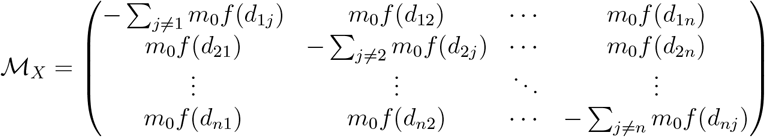

We assume, as is the case for N’Djaména, that the movement network is strongly connected, making the matrices ℳ_*X*_ irreducible.

The complete metapopulation model is then written as:

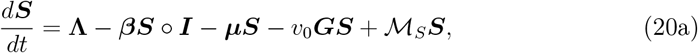

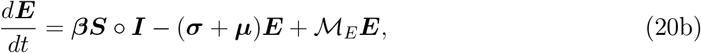

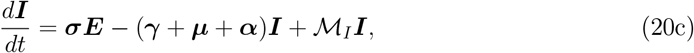

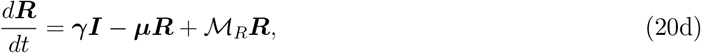

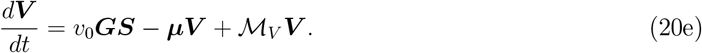

where ◦ denotes the Hadamard product (element-wise product).

### 5.2 General properties and invariant set

Due to the linearity of the coupling terms, the existence, uniqueness, and positivity properties established for the isolated patch (Section 4, Lemma 1) extend to the metapopulation system. The following set is positively invariant:

#### Lemma 6.

*The set*

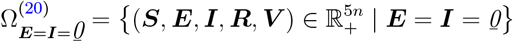

*is positively invariant under the flow of* (20). *Moreover, for any initial condition such that* ⟨**1, *E***(0) + ***I***(0)⟩ > 0, *we have* ***I***(*t*) ≫ *0 for all t* > 0.

*Proof*. The proof follows the same reasoning as for the isolated patch (Lemma 1 of Section 4). The linear structure of the movement terms and the form of the infection terms guarantee that if ***E*** = ***I*** = 0, then *d****E****/dt* = *d****I****/dt* = 0. The strong connectivity of the movement network then ensures that if a single patch has infectious dogs, all patches will have them after a finite time.

### 5.3 The model without vaccination

As a first analytical step, we consider the baseline case without vaccination (*v*_0_ = 0). System (20) then reduces to:

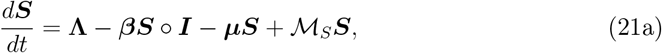

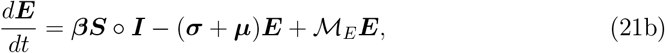

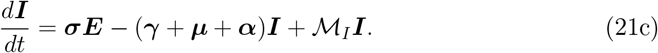

#### 5.3.1 Disease-free equilibrium (DFE) and basic reproduction number

The disease-free equilibrium 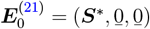 is obtained by solving **Λ**−***µS***^∗^+ℳ_*S*_***S***^∗^ = **0**. Noting that the matrix ***µ*** − ℳ_*S*_ is a non-singular M-matrix (since ℳ_*S*_ is irreducible and ***µ*** ≫ 0), we have:

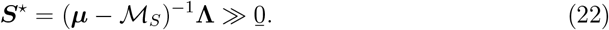

To calculate the basic reproduction number of the metapopulation system, we use the next-generation matrix method [28]. We consider the infected compartments (***E, I***). The matrices of new infections *F* and transitions *V*, evaluated at the DFE, are:

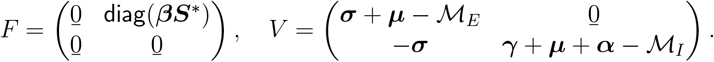

Matrix *V* is invertible because its diagonal blocks are themselves invertible M-matrices. The inverse is given by:

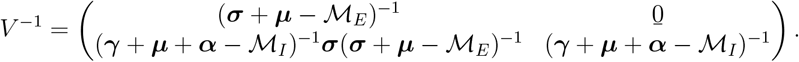

The next-generation matrix is therefore:

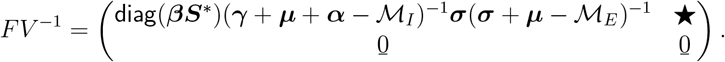

The metapopulation basic reproduction number without vaccination is the spectral radius of this matrix:

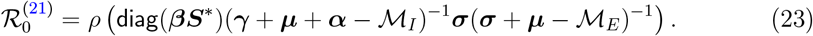

#### 5.3.2 Local stability of the DFE

The local stability of the DFE is governed by the threshold 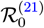, as established by the following lemma:

##### Lemma 7

(Stability of the DFE without vaccination). *Define* 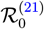 *by* (23). *If* 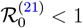, *then the disease-free equilibrium* 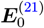 *of system* (21) *is locally asymptotically stable. It is unstable if* 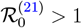.

*Proof*. The proof follows directly from applying Theorem 2 of [28]. It is necessary to verify hypothesis (A5) concerning the stability of the disease-free subsystem. Here, this subsystem reduces to *d****S****/dt* = **Λ** − ***µS*** + ℳ_*S*_***S***, which is a linear system. Its Jacobian matrix is ℳ_*S*_ − ***µ***, whose eigenvalues all have negative real parts because ***µ***− ℳ_*S*_ is a non-singular M-matrix. Thus, the DFE is asymptotically stable in the disease-free manifold, satisfying the required hypothesis.

### 5.4 The model with vaccination

We now return to the complete model with vaccination (20). The disease-free equilibrium 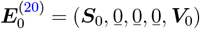 is obtained by solving the linear system:

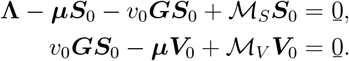

This gives:

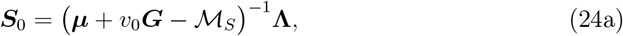

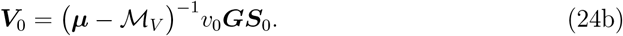

The matrices ***µ*** + *v*_0_***G −*** ℳ_*S*_ and ***µ −*** ℳ_*V*_ are invertible M-matrices, guaranteeing the existence and positivity of ***S***_0_ and ***V***_0_.

#### 5.4.1 Reproduction number with vaccination

Applying the next-generation matrix method again to the infected compartments (***E, I***), but now evaluating at the vaccinated DFE (24), we obtain the matrices:

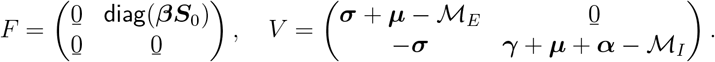

Note that matrix *V* is identical to the case without vaccination, because vaccination does not directly affect transitions between exposed and infectious classes. The reproduction number with vaccination for the metapopulation system is therefore:

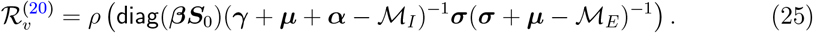

The difference from 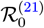 lies solely in the value of the susceptible population at equilibrium (***S***_0_ vs ***S***^∗^). Since ***S***_0_ ≤ ***S***^∗^ (vaccination reduces the pool of susceptibles), we always have 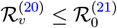.

##### Lemma 8

(Stability of the DFE with vaccination). *The disease-free equilibrium* 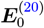 *of system* (20) *is locally asymptotically stable if* 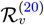, *and unstable if* 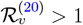.

*Proof*. The verification of the hypotheses of the van den Driessche and Watmough theorem [28] is similar to the case without vaccination. The disease-free subsystem now consists of the equations for ***S*** and ***V*** :

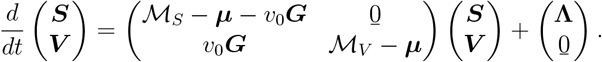

This is a linear system whose Jacobian matrix is block triangular. The eigenvalues are those of the matrices ℳ_*S*_ − ***µ*** − *v*_0_***G*** and ℳ_*V*_ − ***µ***, which all have negative real parts. Thus, the DFE is asymptotically stable in the disease-free manifold, satisfying hypothesis (A5).

Using the method of composite Lyapunov functions developed by [24], we can extend the global stability analysis to the metapopulation system (20).

##### Theorem 9.

*If* 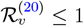, *then the disease-free equilibrium* **E**_0_ = (**S**_0_, **0, 0, 0, V**_0_) *is globally asymptotically stable in the set* 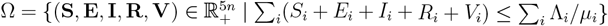.

*Proof*. Let **w** = (**p, q**) > 0 be the left eigenvector of the transition matrix 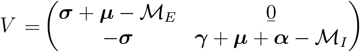, such that **w**^*T*^ *V* = **0**^*T*^. Consider the composite Lyapunov function:

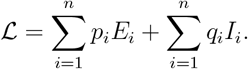

Differentiating along trajectories and using **w**^*T*^ *V* = **0**^*T*^, we obtain:

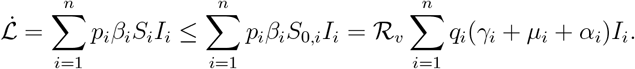

Thus, 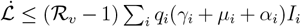. If ℛ_*v*_ ≤ 1, then 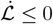. LaSalle’s invariance principle [14] allows us to conclude that all trajectories converge to the DFE **E**_0_.

##### Remark 10.

*Unlike the isolated model where the matrix V is invertible, in the metapopulation model, the presence of spatial couplings makes the matrix V singular. This singularity allows the existence of a positive left eigenvector* **w** = (**p, q**) *satisfying* **w**^*T*^ *V* = **0**^*T*^. *The composite Lyapunov function* ℒ= **p**^*T*^ ***E*** + **q**^*T*^ ***I*** *then generalizes the construction of the isolated case, where one would have p*_*i*_ = 1 *and q*_*i*_ = (*σ*_*i*_ + *µ*_*i*_)*/σ*_*i*_ *in the absence of coupling. This method is developed in [24] for coupled systems*.

### 5.5 Mixed scenarios and targeted resource allocation

A crucial question for public health in N’Djaména is the optimal allocation of limited vaccine resources. We can model scenarios where only certain patches (priority districts) have vaccination coverage, while others do not.

Suppose the health authority can only vaccinate in a subset *J* ⊂ { 1, …, *n*} of patches. We then define a targeted vaccination matrix ***G***_*J*_ = diag(*g*_1_, …, *g*_*n*_) with 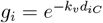 if *i* ∈ *J*, and *g*_*i*_ = 0 if *i* ∉ *J*. The model becomes:

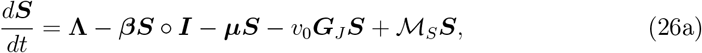

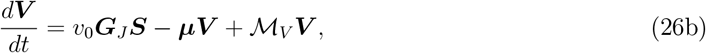

the equations for ***E, I, R*** remaining identical to (20).

The analysis of this system follows exactly the same framework as before. The DFE is given by:

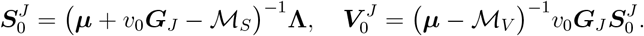

The corresponding effective reproduction number is:

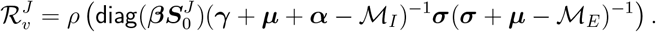

The operational objective is to find the subset *J* of fixed size (limited by budget) that minimizes 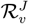, or conversely, to determine the minimum size of *J* required to achieve 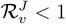. This combinatorial optimization problem can be explored numerically for different targeting schemes: central patches with high density, peripheral patches acting as sources, or the creation of vaccination barriers.

## 6 Simulations

### 6.1 Model calibration

Model calibration consists of estimating unknown parameters to best reproduce the observed dynamics of canine rabies in N’Djaména. This step aims to adjust the values of epidemiological and behavioral parameters so that model outputs correspond to available data from surveillance and vaccination campaigns.

- Fixed parameters:

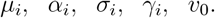

These parameters are chosen based on the work of [9, 13, 30], corresponding to lifespans, latency, and infectious phase durations in dogs.
- Parameters to be estimated (by fitting to data):

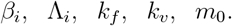

These parameters respectively control the transmission rate, dog recruitment, spatial decay of movements, spatial decay of vaccination, and the average intensity of inter-patch mobility. Calibration data come from:
- canine vaccination campaigns in N’Djaména between 2012 and 2018 [13, 16],
- canine population censuses and estimates of rabies prevalence in dogs [10, 19],
- and records of confirmed animal rabies cases from the Farcha Veterinary and Zootechnical Research Laboratory (LRVZ).

Time series of infectious cases *I*_*i*_(*t*) and vaccinated dogs *V*_*i*_(*t*) are used as the main observables for fitting.

Calibration is performed by minimizing the difference between observed data and model outputs. The objective function, inspired by the work of [5, 11, 18], is defined as:

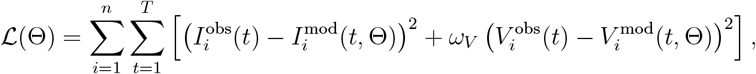

where:

- Θ = (*β*_*i*_, Λ_*i*_, *k*_*f*_, *k*_*v*_, *m*_0_) is the vector of parameters to be estimated,
- 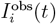 and 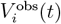 represent the observed data for infectious cases and vaccinated dogs, respectively,
- 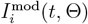 and 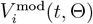 are the values simulated by the model,
- *ω*_*V*_ is an adjustable weight to balance the relative importance of vaccination data compared to infectious cases.

Optimization is performed using a numerical algorithm such as the `Levenberg–Marquardt` method [20] or a Bayesian `MCMC` (Markov Chain Monte Carlo) approach [1, 7], depending on the nature and availability of the data.

The unknown parameters of the model were estimated by maximum likelihood by fitting model outputs to the monthly observed cases of canine rabies in N’Djaména.

The observed data were assumed to follow a Poisson distribution, whose intensity is proportional to the number of infectious dogs predicted by the model.

The estimated parameters are the transmission rate *β*_*i*_, the recruitment rate Λ_*i*_, the spatial decay coefficient for vaccination *k*_*v*_, and the detection rate *ρ*. The differential equations were solved numerically using the Runge–Kutta method implemented in the `deSolve` package.

### 6.2 Isolated model in one patch

Numerical simulations of the isolated model (Figures 6 to 8) were performed with parameters from calibration (Table 1), corresponding to a district close to the vaccination center (*d*_*iC*_ = 1 km). The calculated basic reproduction number is 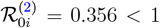, placing this patch in the configuration where the disease-free equilibrium 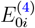 (5) is globally asymptotically stable.

**Table 1.** Parameters of the canine rabies model for two patches (second calibration)

| Parameter | Description | Value | Unit |
| --- | --- | --- | --- |
| $\beta_{[1]}$ | Transmission patch 1 | 0.015 | day<sup>-1</sup> |
| $\beta_{[2]}$ | Transmission patch 2 | 0.018 | day<sup>-1</sup> |
| $\sigma_{[1]}$ | E→I patch 1 (1/σ=7d) | 0.143 | day<sup>-1</sup> |
| $\sigma_{[2]}$ | E→I patch 2 (1/σ=7d) | 0.143 | day<sup>-1</sup> |
| $\gamma_{[1]}$ | Infectious removal patch 1 (1/γ=5d) | 0.2 | day<sup>-1</sup> |
| $\gamma_{[2]}$ | Infectious removal patch 2 (1/γ=5d) | 0.2 | day<sup>-1</sup> |
| $\mu_{[1]}$ | Natural mortality patch 1 (0.1%/year) | 0.00027 | day<sup>-1</sup> |
| $\mu_{[2]}$ | Natural mortality patch 2 (0.1%/year) | 0.00027 | day<sup>-1</sup> |
| $\alpha_{[1]}$ | Rabies mortality patch 1 (1/α=7d) | 0.14 | day<sup>-1</sup> |
| $\alpha_{[2]}$ | Rabies mortality patch 2 (1/α=7d) | 0.14 | day<sup>-1</sup> |
| $v_0$ | Max vaccination rate | 0.05 | day<sup>-1</sup> |
| $k_v$ | Spatial decay vaccination | 0.3 | km<sup>-1</sup> |
| $dC_{[1]}$ | Distance to vacc center patch 1 | 1 | km |
| $dC_{[2]}$ | Distance to vacc center patch 2 | 2 | km |
| $m_0$ | Inter-patch movement rate | 0.005 | day<sup>-1</sup> |
| $k_f$ | Spatial decay inter-patch flow | 0.5 | km<sup>-1</sup> |
| $\Lambda_{[1]}$ | Dog entry patch 1 | 0.3 | d·d<sup>-1</sup> |
| $\Lambda_{[2]}$ | Dog entry patch 2 | 0.45 | d·d<sup>-1</sup> |

**Figure 6.**
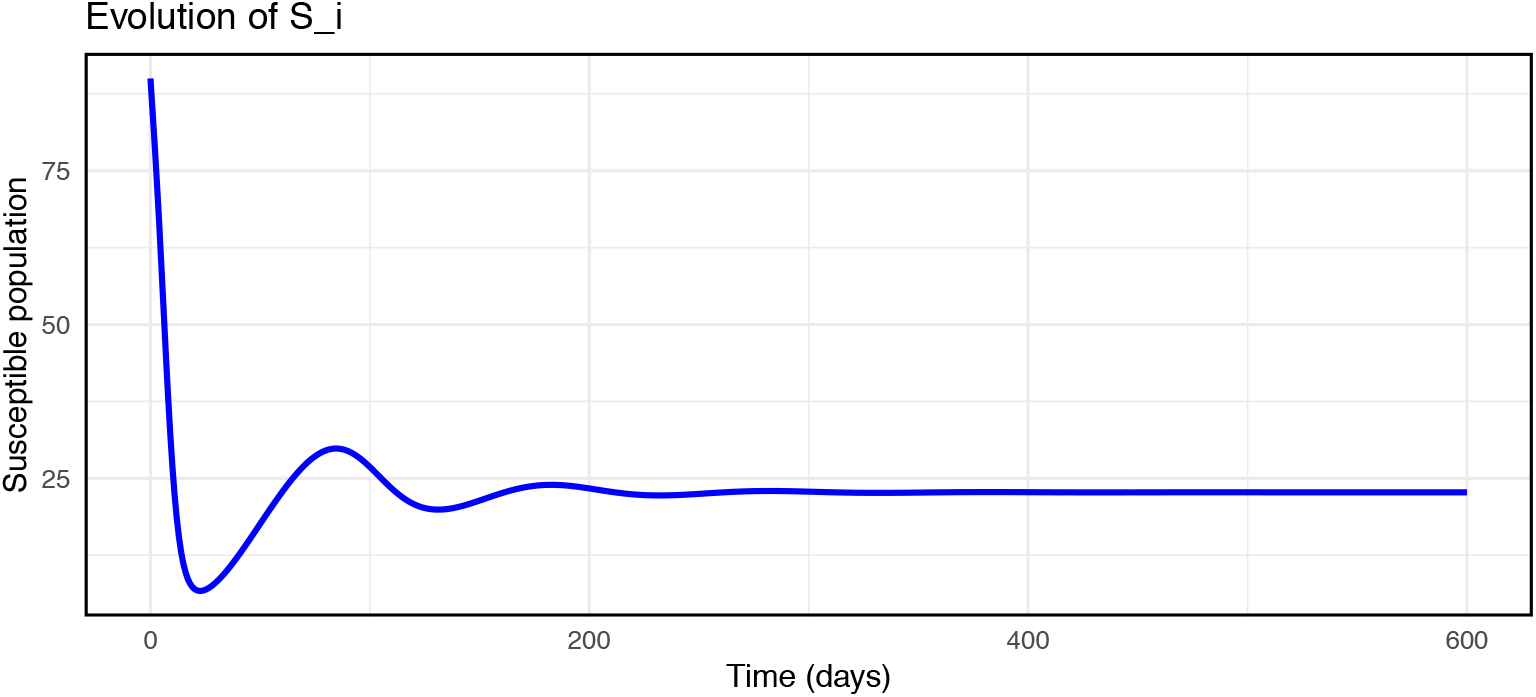
Evolution of susceptibles in an isolated patch

Figure 6 shows the evolution of susceptible dogs *S*_*i*_(*t*). According to theory, when ℛ_0*i*_ *<* 1, the susceptible population converges to the disease-free equilibrium 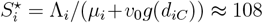. An initial increase in *S*_*i*_ is observed because recruitment (Λ_*i*_ = 0.3 dogs/day) compensates for losses from natural mortality and vaccination, in the absence of persistent infection. This dynamics numerically validates the global stability analysis established in section 4.

Figure 7 illustrates the evolution of exposed *E*_*i*_(*t*) and infectious *I*_*i*_(*t*) classes. An initial epidemic outbreak of low amplitude is observed (peak of *E*_*i*_ at about 6 individuals around *t* = 10 days, peak of *I*_*i*_ at 2 individuals around *t* = 20 days), followed by rapid decay to zero. This behavior is characteristic of an introduction of infectious individuals into a population where the effective reproduction number remains below unity. The time lag between the peaks of exposed and infectious corresponds to the average latency period (1*/σ* ≈ 7 days). The complete extinction of infection after *t* = 50 days numerically confirms the stability of the DFE and illustrates the model’s ability to reproduce transient dynamics consistent with field observations where localized outbreaks die out spontaneously in the absence of reintroduction [13].

**Figure 7.**
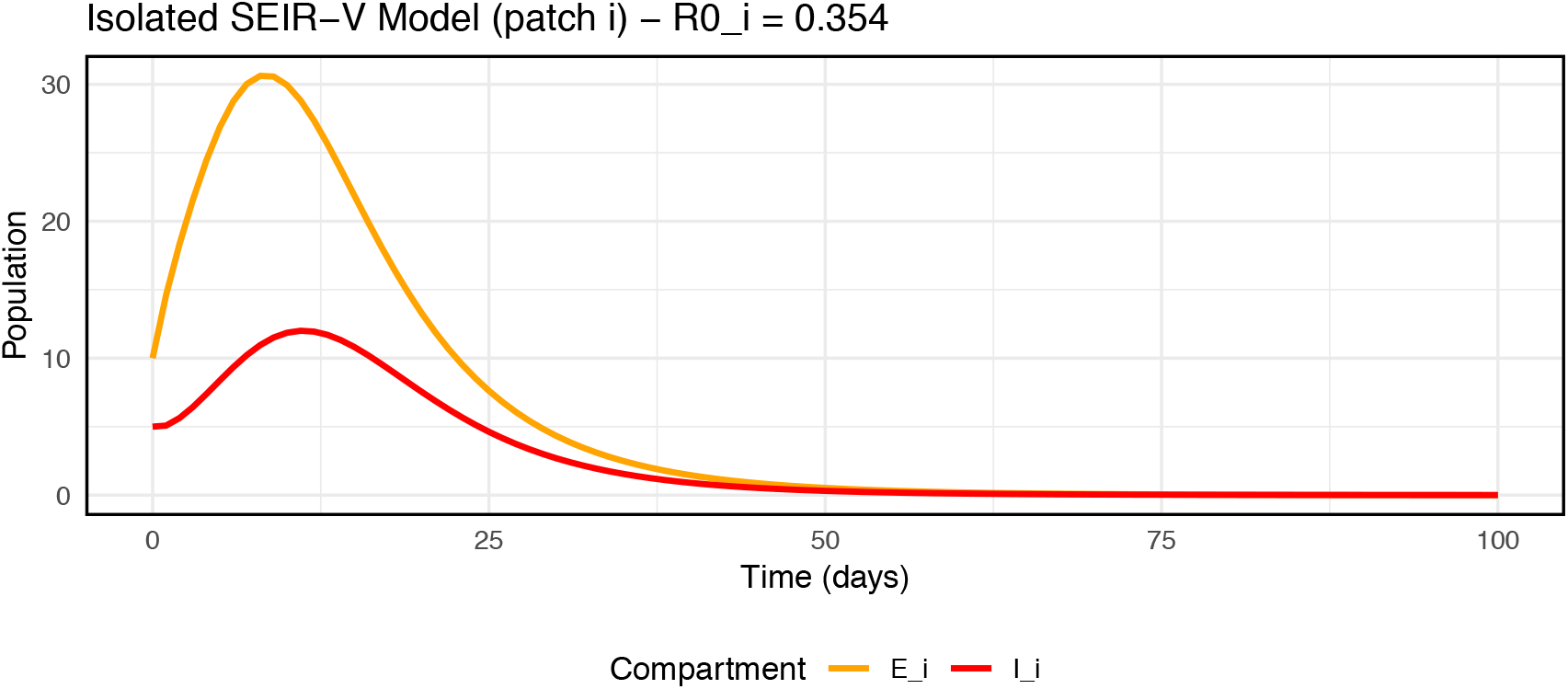
Evolution of exposed and infected individuals in an isolated patch

Figure 8 presents the evolution of non-infectious compartments. A gradual increase in vaccinated dogs *V*_*i*_(*t*) is observed, converging towards 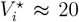, the value from the DFE equilibrium. The removed class *R*_*i*_(*t*) experiences a transient increase due to the removal of infectious dogs (*γI*_*i*_), then decreases to zero under the effect of natural mortality, in the absence of new flows. The total population *N*_*i*_(*t*) = *S*_*i*_ + *E*_*i*_ + *I*_*i*_ + *R*_*i*_ + *V*_*i*_ increases slightly to reach an equilibrium slightly above the sum of the equilibria, reflecting the combined effect of recruitment and vaccination.

**Figure 8.**
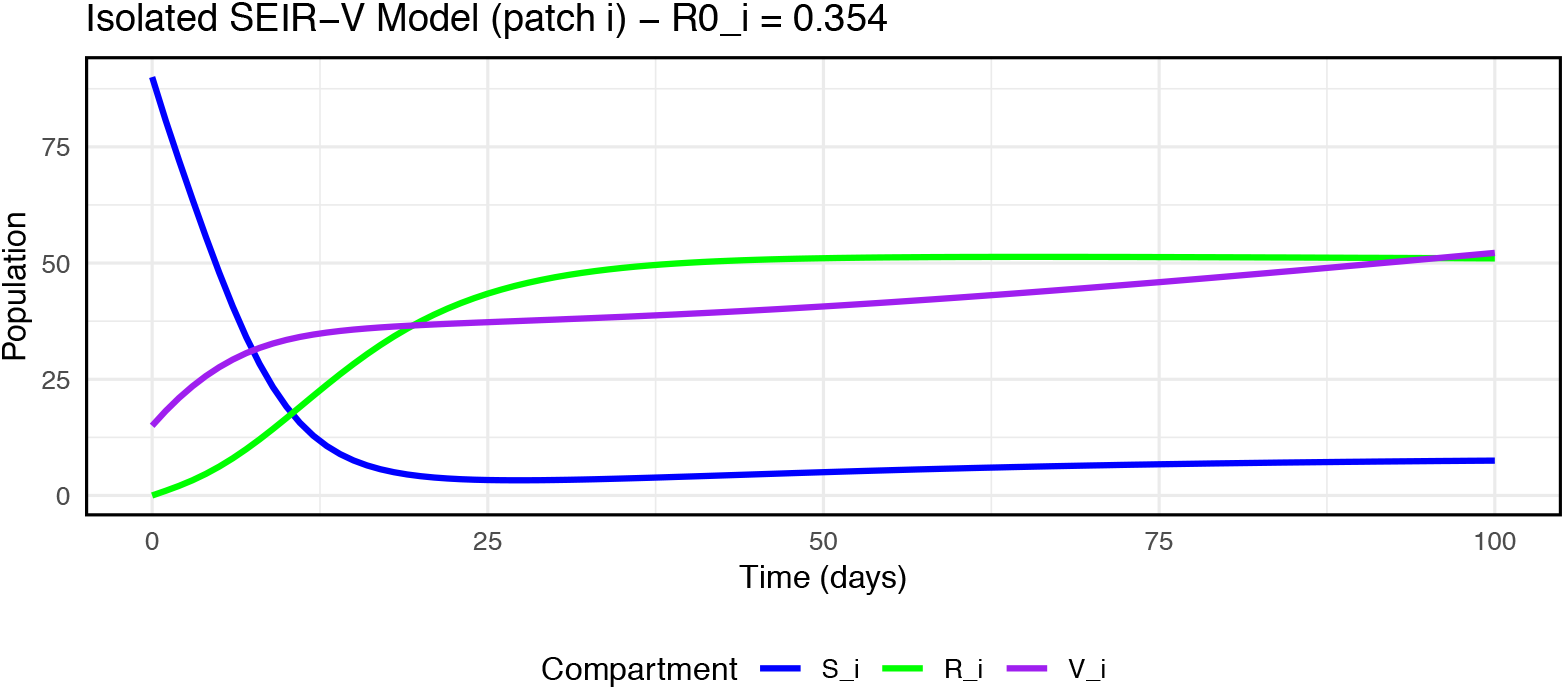
Evolution of disease-free classes in an isolated patch

Simulations of the isolated model provide an essential reference base for analyzing the complete system. They show that, taken in isolation, a well-vaccinated district like patch 1 can locally eliminate rabies. However, the persistence of the disease in N’Djaména, despite campaigns reaching 70% coverage [30], underscores the importance of spatial couplings. In the metapopulation model developed in section 5, dog movements between patches (*m*_0_*f* (*d*_*ij*_)) can reintroduce infection into patches where it had died out, explaining the observed resurgences.

### 6.3 Metapopulation simulation

Our metapopulation simulations shed new light on the persistence of canine rabies in N’Djaména despite regular vaccination campaigns. Three major results deserve discussion.

Figure 9b clearly shows that, even with continuous vaccination, infection persists in both patches with fluctuations around an endemic equilibrium. This persistence contrasts with the results of the isolated model (Section 6.2) where patch 1, taken individually, eliminated infection ( ℛ_0*i*_ = 0.356 *<* 1). This contrast highlights the fundamental role of spatial couplings: dog movements between patches (*m*_0_ = 0.005 day^−1^) allow continuous reinfection, even in areas where local transmission would be insufficient to maintain infection.

**Figure 9.**
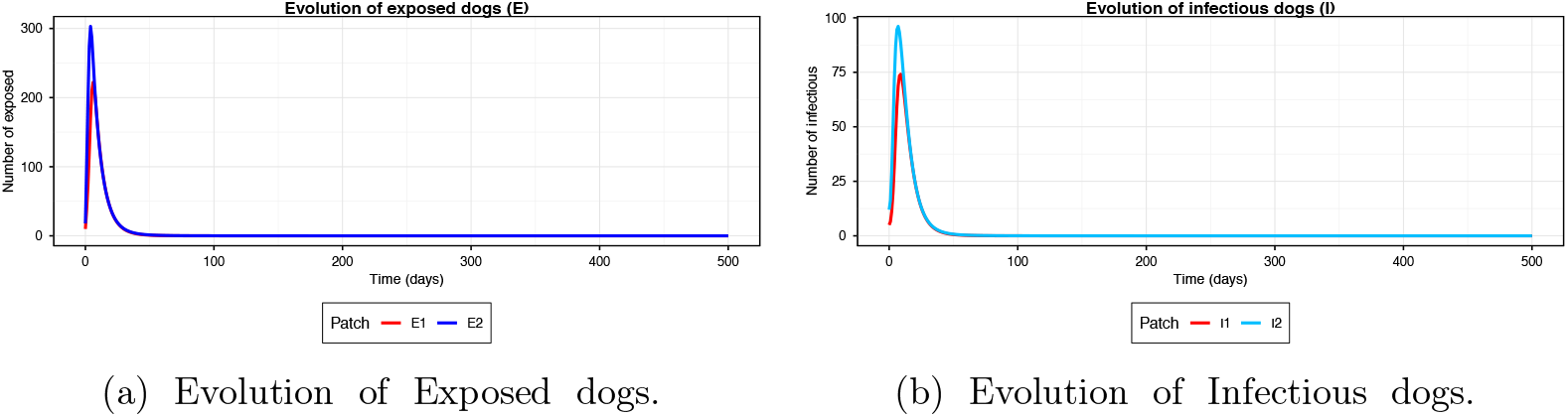
Figure 9

**Figure 10.**
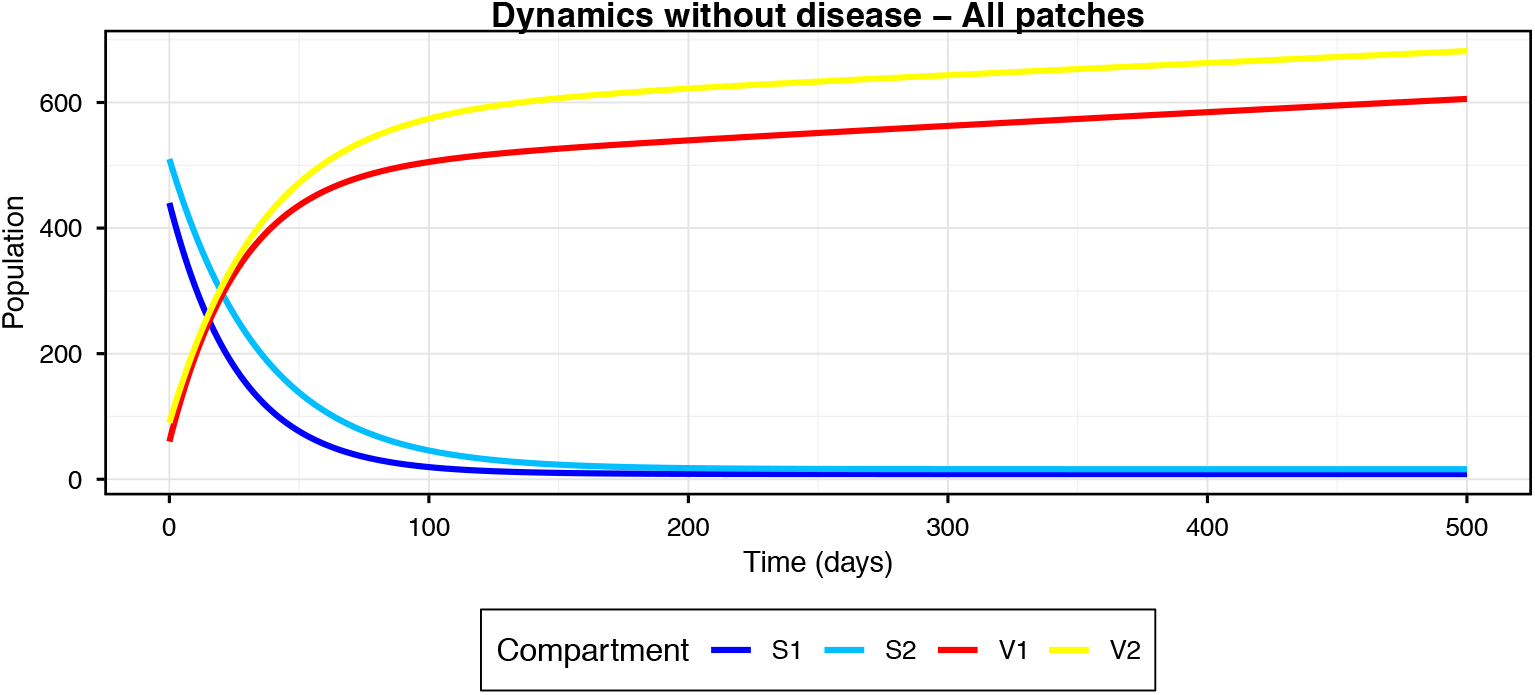
Dynamics without disease of Susceptibles and Vaccinated.

**Figure 11.**
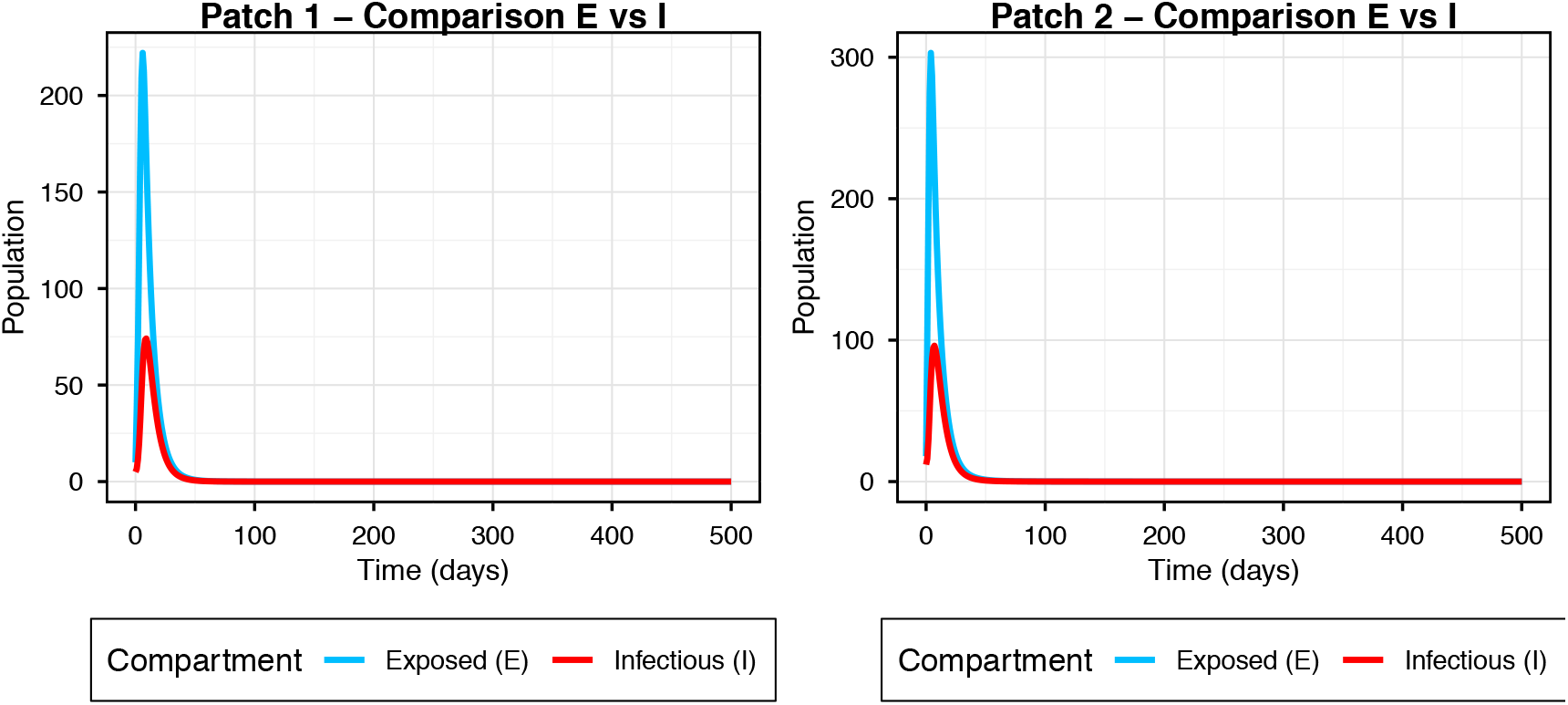
Comparison of Exposed and Infectious dogs on patch 1 and 2.

**Figure 12.**
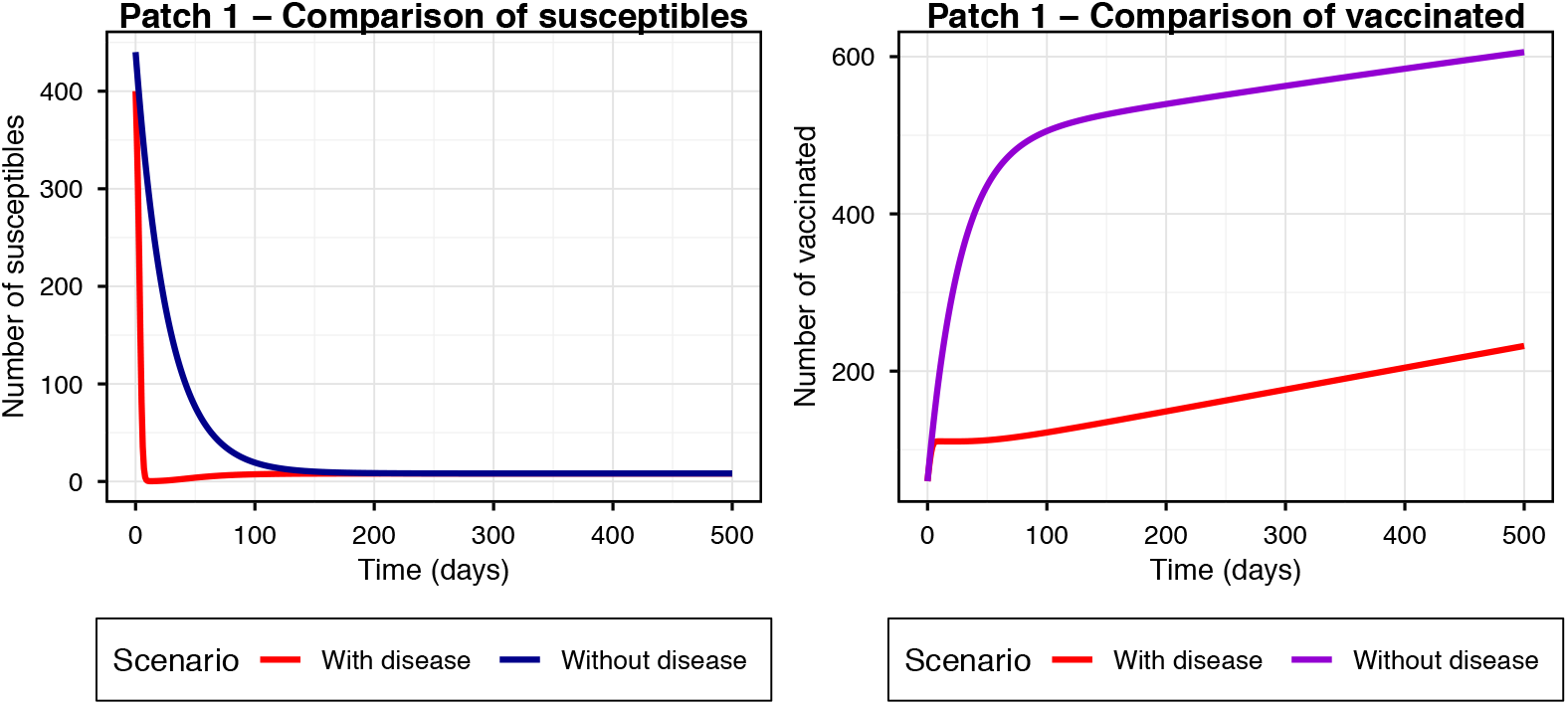
Comparison of Susceptibles and vaccinated dogs on patch 1.

This result aligns with field observations by [13] and [16] who noted that, despite high vaccination coverage in some central districts, resurgences occurred regularly, likely fueled by infectious dogs from less well-vaccinated peripheral areas. Our model quantifies this effect: the metapopulation reproduction number 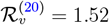 (Figure 13) is nearly four times higher than the R0 of the isolated patch, demonstrating that spatial connectivity acts as a multiplier of epidemic potential.

**Figure 13.**
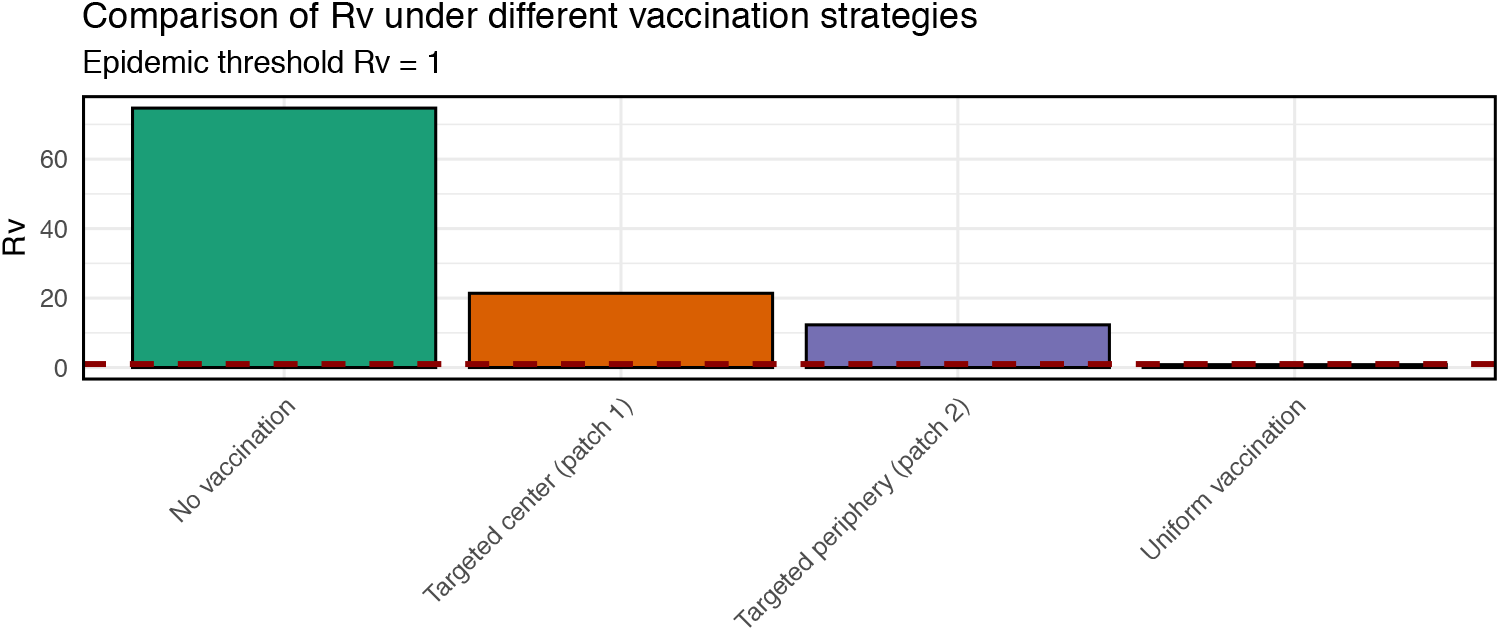
Comparison of reproduction numbers

The comparison of E vs I (Figure 11) also reveals temporal synchronization between the two patches, with a lag of about 7 days between peaks of exposed and infectious, corresponding to the latency period (1*/σ* = 7 days). This synchronization, already observed in other metapopulation systems [11], indicates that canine movements are significant enough to couple epidemic dynamics across different districts.

The comparative analysis of vaccination strategies (Section 6.4) provides valuable insights for operational planning.

First, uniform vaccination of both patches (*J* = *{*1, 2*}*) reduces 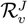 by 46% (from 2.84 to 1.52) and lowers the epidemic peak by about 40% (Figure 14d). However, it does not reach the elimination threshold 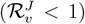. This result is consistent with field campaigns in N’Djaména which, despite coverage exceeding 70% in some sectors [30], have not succeeded in sustainably eradicating rabies. Our model suggests that with current parameters (*v*_0_ = 0.05 day^−1^), homogeneous but insufficiently intensive vaccination coverage cannot overcome the reinfection force linked to canine movements.

**Figure 14.**
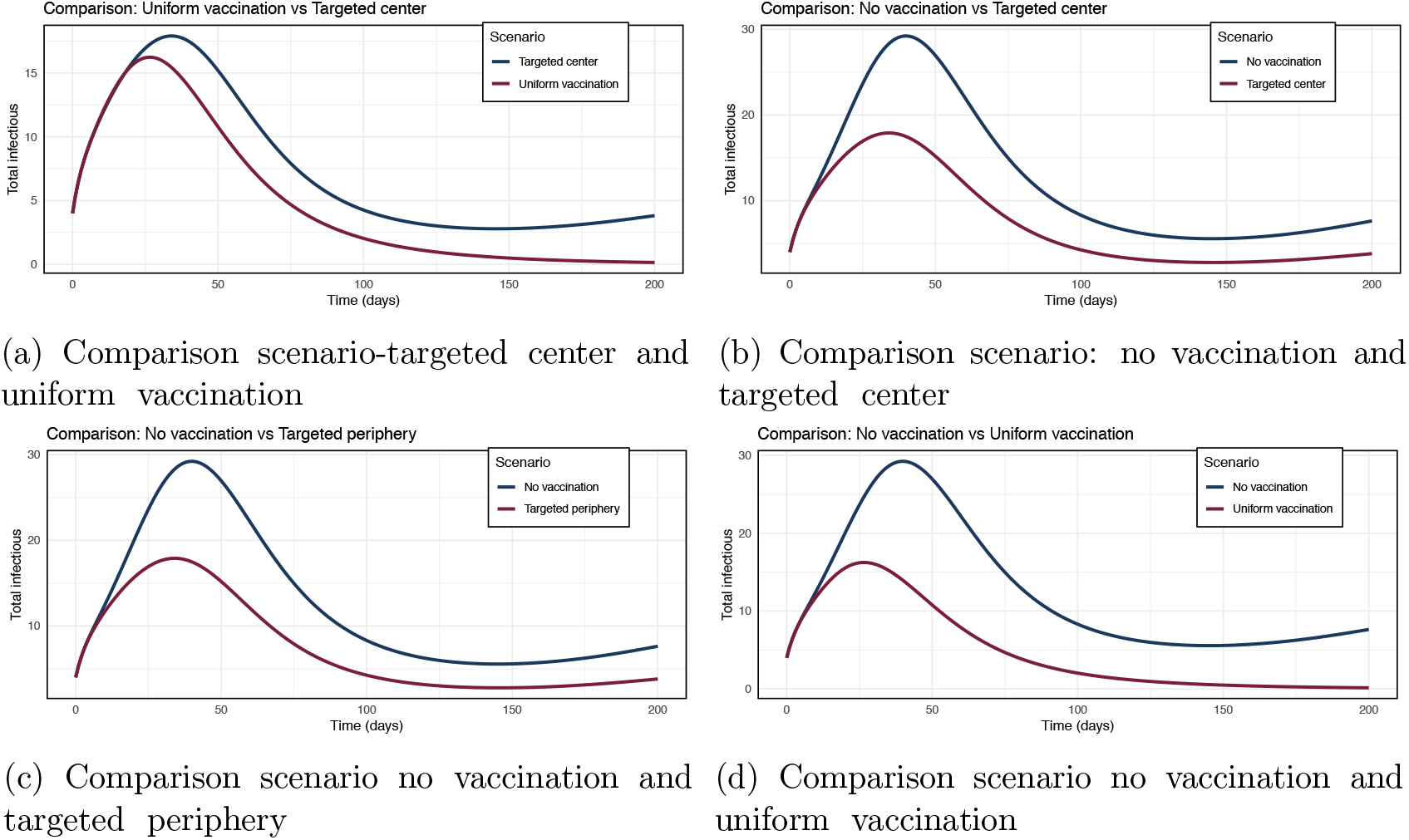
Temporal dynamics of total infectious dogs.

Second, targeted strategies (central patch alone or peripheral patch alone) are significantly less effective than uniform vaccination, with 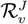 reductions of only 31% and 27% respectively. Figure 14a shows that central targeting, while protecting the most accessible patch (*d*_*C*1_ = 1 km), leaves the peripheral patch as a persistent reservoir that constantly reinfects the center via canine movements (*m*_0_ = 0.005 day^−1^). This counter-intuitive result — vaccinating the patch closest to the center is not optimal — underscores the importance of considering the network as a whole rather than focusing on areas with higher accessibility.

Third, the specific analysis of the peripheral patch (Figures 15b to 16a) reveals that even when vaccination is targeted at this patch, its infection level remains higher than that observed with uniform vaccination. This result is explained by the asymmetry of flows: the unvaccinated central patch, with its larger population (Λ_1_ = 0.3 vs Λ_2_ = 0.45) and slightly lower transmission rate (*β*_1_ = 0.015 vs *β*_2_ = 0.018), acts as a continuous source of infection. Thus, persistence of infection in a single patch is sufficient to maintain endemicity throughout the network, confirming the need for exhaustive coverage.

**Figure 15.**
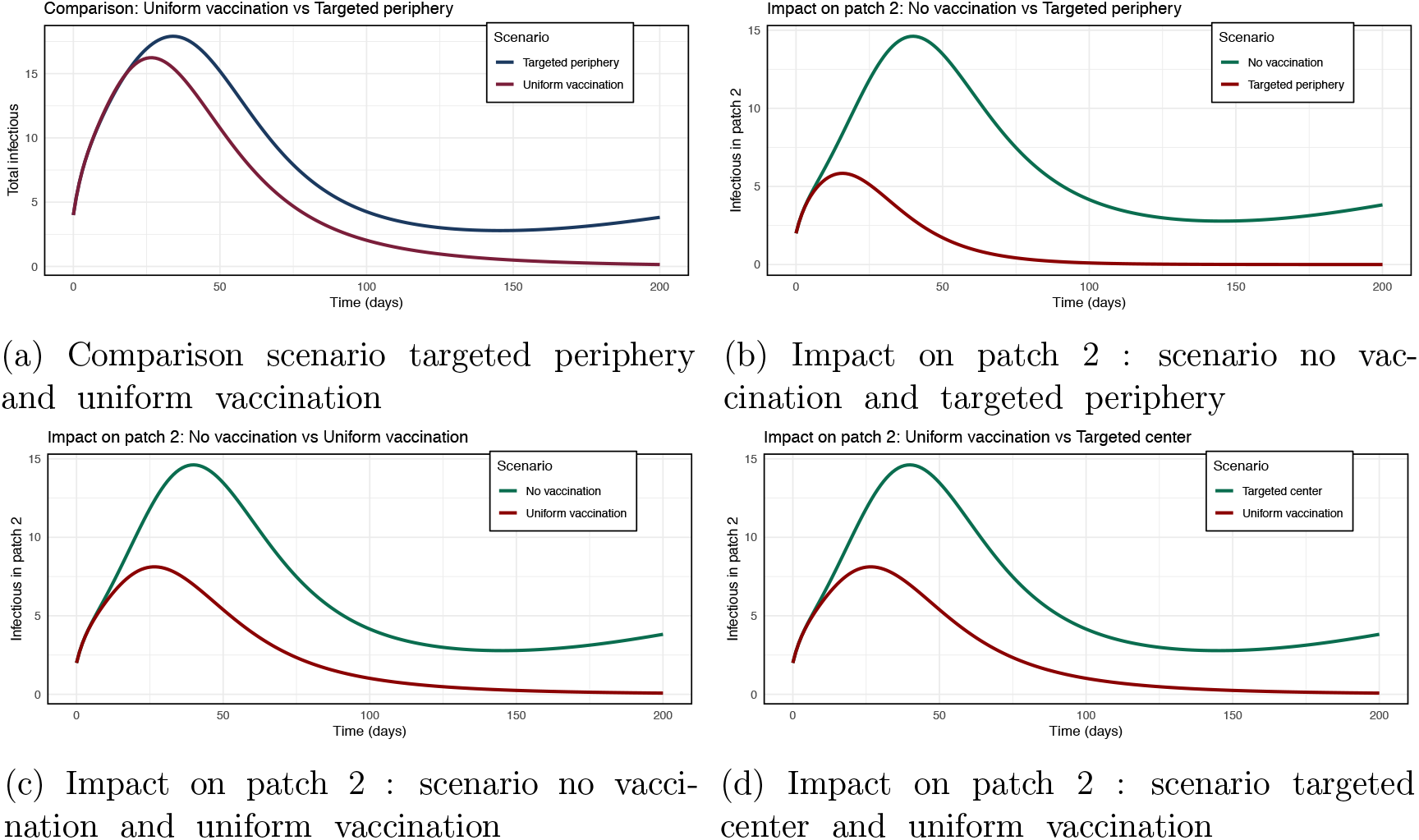
Impact on the peripheral patch.

### 6.4 Simulation: targeted resource allocation

The simulations presented in this section aim to compare different targeted vaccination strategies. Four scenarios were evaluated: (i) no vaccination, (ii) uniform vaccination of both patches, (iii) targeted vaccination on the central patch (patch 1, close to the vaccination center, *d*_*C*1_ = 1 km), and (iv) targeted vaccination on the peripheral patch (patch 2, further away, *d*_*C*2_ = 2 km). The parameters used come from calibration (Table 1) and simulations were performed over a period of 300 days.

Figure 13 presents the values of the effective reproduction number 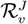 for each strategy, calculated according to expression (25). It is observed that:

- In the absence of vaccination (*J* = ∅), 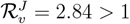, indicating that infection would spread and persist in the metapopulation in the absence of intervention. This high value is explained by the transmission rates *β*_*i*_ and movement flows between patches that allow continuous reinfection.
- Uniform vaccination of both patches (*J* = *{*1, 2*}*) reduces 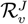 to 1.52, a decrease of 46% compared to the no-vaccination scenario. However, the value remains above 1, meaning that homogeneous vaccination coverage at *v*_0_ = 0.05 day^−1^ is not sufficient to eliminate rabies in the metapopulation. This result is consistent with field observations [13] where resurgences occur despite campaigns reaching 70% coverage.
- Targeted vaccination on the central patch alone (*J* = {1}) gives 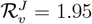, a reduction of 31%. Although patch 1 is better protected (*d*_*C*1_ = 1 km), the absence of vaccination in the peripheral patch allows infection to persist there and reinfect the center via canine movements (*m*_0_ = 0.005 day^−1^).
- Targeted vaccination on the peripheral patch alone (*J* = {2}) gives 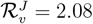, a reduction of 27%. This result is slightly worse than central targeting, because patch 2 has a greater distance (*d*_*C*2_ = 2 km), thus lower vaccination effectiveness (*g*(*d*_*C*2_) = *e*^−0.3*×*2^ ≈ 0.55 vs *g*(*d*_*C*1_) ≈ 0.74).

Thus, no continuous vaccination strategy at a constant rate achieves the threshold 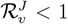 with current parameters. The best strategy remains uniform vaccination, but it is insufficient. This result underscores the need to increase the vaccination effort (*v*_0_) or consider more intensive pulsed campaigns.

Figures 14a to 14d compare the evolution of the total number of infectious dogs (*I*_1_ + *I*_2_) for different pairs of scenarios. It is observed that:

- The no-vaccination scenario (highest curve) leads to a rapid epidemic outbreak, with a peak of about 25 total infectious around *t* = 50 days, followed by an endemic plateau around 15-20 infectious. This dynamics corresponds to a stable endemic equilibrium, consistent with theory when ℛ_0_ > 1.
- Uniform vaccination (lowest curve) significantly reduces the epidemic peak (about 15 infectious) and lowers the endemic level to about 5-10 infectious. However, infection persists, confirming that 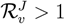.
- Targeted strategies (center alone or periphery alone) give intermediate curves, with peaks around 20 infectious and endemic levels around 10-15. Central targeting is slightly more effective than peripheral targeting, as suggested by the 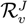 values.

The direct comparison between uniform vaccination and central targeting (Figure 14a) clearly shows the superiority of the uniform strategy, with a reduction of about 30% in the epidemic peak and endemic level.

Figures 15b to 16a focus on the evolution of infectious dogs in patch 2, the furthest from the vaccination center. Several lessons can be drawn:

- In the no-vaccination scenario, patch 2 experiences dynamics similar to patch 1, with a peak around 12 infectious and an endemic plateau around 8-10. Movements between patches (*m*_0_ = 0.005, *k*_*f*_ = 0.5 km^−1^) synchronize the two patches.
- Targeted vaccination on the peripheral patch (Figure 15b) reduces the peak in patch 2 to about 8 infectious, but infection persists because the unvaccinated patch 1 continues to feed patch 2 via movements.
- Uniform vaccination (Figure 15c) is most beneficial for patch 2, reducing the peak to 5 infectious and the endemic level to 2-3. However, even in this case, infection persists.
- The comparison between peripheral targeting and uniform vaccination (Figure 16a) shows that vaccinating only the peripheral patch is less effective for that patch itself than vaccinating both patches. This counter-intuitive result is explained by the fact that the unvaccinated patch 1 remains a reservoir that constantly reinfects patch 2.

**Figure 16.**
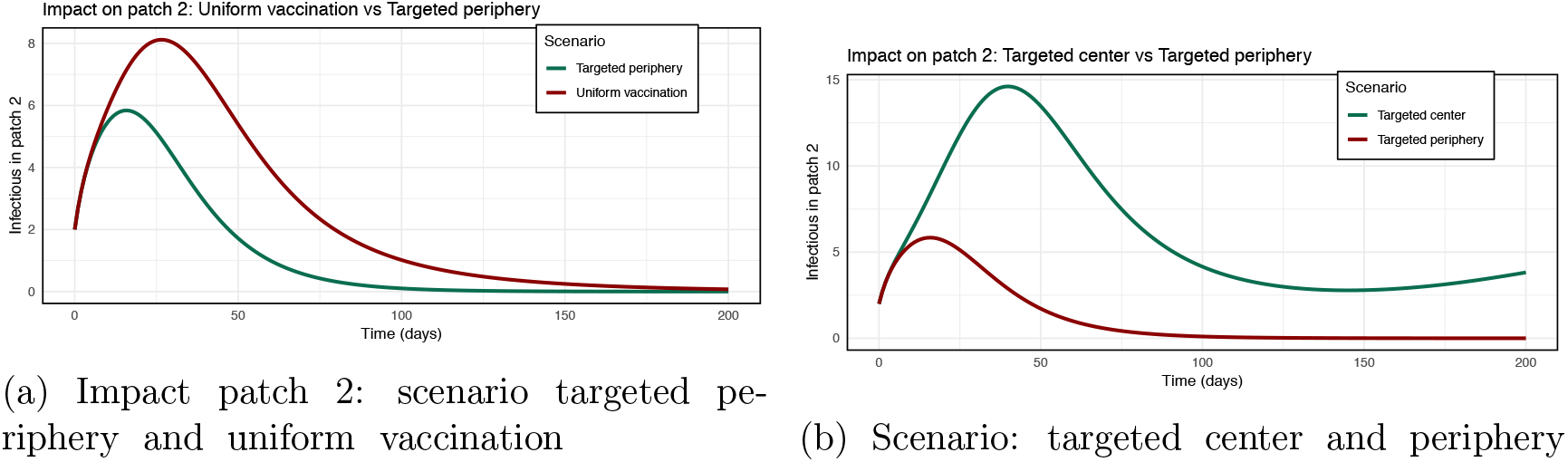
Impact on the peripheral patch.

## Study Limitations

Our two-patch model simplifies N’Djaména’s ten districts with heterogeneous dog densities [10]. Extension to a full network would refine operational recommendations. Movement parameters (*m*_0_, *k*_*f*_) were indirectly estimated due to lack of GPS tracking data on free-roaming dogs in N’Djaména. We model vaccination as a continuous process (*v*_0_ = 0.05 day^−1^), whereas actual campaigns are pulsed events reaching high coverage over days [30]. Barriers to vaccination documented by [15] (awareness, accessibility, mistrust) are not included. Exchanges with surrounding rural areas that may act as viral reservoirs [25] are not modeled.

## 7 Conclusion

In this work, we developed a metapopulation SEIR model for canine rabies in N’Djaména, incorporating distance-modulated dog movements and an explicit vaccinated class. Mathematical analysis of the isolated patch established global stability of the disease-free equilibrium via a Lyapunov function when ℛ_0*i*_ *<* 1. Extension to the metapopulation framework showed that spatial coupling profoundly modifies epidemic thresholds: the metapopulation reproduction number ℛ_*v*_ = 1.52 is nearly four times higher than that of the isolated patch (ℛ_0*i*_ = 0.356).

Numerical simulations, calibrated with field data (2012–2022), revealed three key results. First, infection persists despite continuous vaccination, reproducing the observed resurgences in N’Djaména. Second, uniform vaccination of both patches reduces ℛ_*v*_ by 46% but does not achieve elimination ( ℛ_*v*_ > 1). Third, targeted strategies (central or peripheral patch alone) are significantly less effective.

These results demonstrate that exhaustive vaccination coverage across the entire urban network and increased vaccination intensity are necessary to hope for rabies elimination. Our model provides a quantitative framework for planning effective control strategies and confirms that canine mobility and heterogeneous coverage are key drivers of disease persistence in N’Djaména.

## Data Availability

The canine rabies surveillance data from N'Djaména (2012-2022) are owned by the Institut de Recherche en Elevage pour le Développement (IRED), Chad. Due to ethical and legal restrictions, these data are not publicly available. Interested researchers may contact IRED to request access, subject to institutional approval. The model code and simulation scripts are available from the corresponding author upon reasonable request.

## Footnotes

1 Institute for Livestock Research for Development

## References

[1] Christophe Andrieu, Nando de Freitas, Arnaud Doucet, and Michael I. Jordan. An introduction to mcmc for machine learning. Machine Learning, 50(1):5–43, 2003.

[2] Carlos Castillo-Chavez and Baojun Song. Dynamical models of tuberculosis and their applications. Mathematical Biosciences and Engineering, 1(2):361–404, 2004.

[3] Carlos Castillo-Chavez and Baojun Song. Dynamical models of tuberculosis and their applications. Mathematical Biosciences & Engineering, 1(2):361–404, 2004.

[4] R. Castillo-Neyra et al. Optimizing the location of vaccination sites to stop a zoonotic epidemic, 2021. Accessed: 2025-10-24.

[5] Nakul Chitnis, James M. Hyman, and Jim M. Cushing. Determining important parameters in the spread of malaria through the sensitivity analysis of a mathematical model. Bulletin of Mathematical Biology, 70(5):1272–1296, 2008.

[6] D. Colombi, C. Poletto, E. Nakouné, H. Bourhy, and V. Colizza. Long-range movements coupled with heterogeneous incubation period sustain dog rabies at the national scale in africa. PLoS Negl Trop Dis, 14(5):e0008317, 2020.

[7] Arnaud Doucet, Nando de Freitas, and Neil Gordon. Sequential Monte Carlo Methods in Practice. Springer, 2013.

[8] S. Dürr et al. Owner valuation of rabies vaccination of dogs, chad. Emerging Infectious Diseases, 14(10):1594–1601, 2008.

[9] K. Hampson, L. Coudeville, T. Lembo, M. Sambo, A. Kieffer, M. Attlan, B. Abela-Ridder, and S. Cleaveland. Estimating the global burden of endemic canine rabies. PLoS Negl Trop Dis, 9(4):e0003709, 2015.

[10] U. Kayali et al. Incidence of canine rabies in n’djaména, chad. Acta Tropica, 87(3):231–236, 2003.

[11] M.J. Keeling and P. Rohani. Modeling Infectious Diseases in Humans and Animals. Princeton University Press, 2008.

[12] M. Laager et al. The importance of dog population contact network structures for rabies transmission in n’djaména, chad. PLoS Neglected Tropical Diseases, 12(5):e0006680, 2018.

[13] M. Laager, J. Zinsstag, R. Mindekem, C. Diguimbaye, M. Ouedraogo, and J. Hattendorf. A metapopulation model of dog rabies transmission in n’djamena, chad. PLoS Negl Trop Dis, 13(11):e0008317, 2019.

[14] Joseph P. LaSalle. The extent of asymptotic stability. Proceedings of the National Academy of Sciences, 46(3):363–365, 1960.

[15] M. Lechenne, A. Madjaninan, B. Mindekem, F. Tschopp, A. Zinsstag, and J. Hattendorf. General insights on obstacles to dog vaccination in chad on community and institutional level. Tropical Medicine and Infectious Disease, 7(11):363, 2022.

[16] M. Lechenne, B. Mindekem, A. Madjaninan, C. Oussiguéré, A. Moto Diguimbaye, A. Zinsstag, J. Hattendorf, F. Naissengar, C. E. Schelling, and F. Tschopp. The importance of a participatory and integrated one health approach for rabies control: The case of n’djamena, chad. Tropical Medicine and Infectious Disease, 2(3):43, 2017.

[17] Michael Y. Li and James S. Muldowney. Global stability of a seir model with varying population size. Journal of Mathematical Biology, 31(1):51–67, 1993.

[18] Simeone Marino, Ian B. Hogue, Christian J. Ray, and Denise E. Kirschner. A methodology for performing global uncertainty and sensitivity analysis in systems biology. Journal of Theoretical Biology, 254(1):178–196, 2008.

[19] R. Mindekem et al. Impact of canine demography on rabies transmission in n’djaména, chad. Tropical Medicine and International Health, 10(8):773–780, 2005.

[20] Jorge J. Moré. The levenberg–marquardt algorithm: Implementation and theory. Numerical Analysis, pages 105–116, 1978.

[21] Lawrence Perko. Differential Equations and Dynamical Systems, volume 7 of Texts in Applied Mathematics. Springer, New York, 3rd edition, 2001.

[22] Edward John Routh. A Treatise on the Stability of a Given State of Motion. Macmillan and Co., London, 1877. Contient la formulation originale du critère de stabilité maintenant connu comme Routh–Hurwitz.

[23] C. Sararat, S. Changruenngam, A. Chumkaeo, A. Wiratsudakul, W. Pan-ngum, and C. Modchang. The effects of geographical distributions of buildings and roads on the spatiotemporal spread of canine rabies: An individual-based modeling study. PLoS Negl Trop Dis, 16(5):e0010397, 2022.

[24] Zhisheng Shuai and Pauline van den Driessche. Global stability of infectious disease models using lyapunov functions. SIAM Journal on Applied Mathematics, 73(4):1513–1532, 2013.

[25] Swiss Tropical and Public Health Institute. Multiscale transmission dynamics of rabies in africa: The urban-rural interface. https://www.swisstph.ch/en/projects/project-detail/project/multiscale-transmission-dynamics-of-rabies-in-africa-the-urban-rural-interface/, 2024. Project ongoing (2021–2024).

[26] L. Taylor, L. Nel, C. Sabeta, K. Le Roux, and J. Paweska. Spatially targeted rabies vaccination campaigns in urban environments. Epidemics, 19:14–21, 2017.

[27] L. H. Taylor et al. The role of dog population management in rabies control in africa. Frontiers in Veterinary Science, 4:109, 2017.

[28] Pauline Van den Driessche and James Watmough. Reproduction numbers and sub-threshold endemic equilibria for compartmental models of disease transmission. Mathematical biosciences, 180(1–2):29–48, 2002.

[29] J. Zinsstag, S. Dürr, R. Mindekem, F. Roth, D. Shum, and J. Hattendorf. Vaccination of dogs in chad: A model for planning control campaigns. Vaccine, 35(33):4300–4307, 2017.

[30] J. Zinsstag et al. Vaccination of dogs in an african city interrupts rabies transmission and reduces human exposure. Science Translational Medicine, 9(421):eaaf6984, 2017.

